# Transcriptomics-Driven Machine Learning Models Accurately Predict Chemotherapy Response in Muscle-invasive Bladder Cancer

**DOI:** 10.1101/2024.06.28.24309634

**Authors:** Ariadna Acedo-Terrades, Alejo Rodriguez-Vida, Oscar Buisan, Marta Bódalo-Torruella, Maria Gabarrós, Miquel Clarós, Nuria Juanpere, Marta Lorenzo, Sergio Vázquez Montes de Oca, Alejandro Rios-Hoyo, Cristina Carrato Moñino, Tamara Sanhueza, Eduardo Eyras, Eulàlia Puigdecanet, Gottfrid Sjödahl, Júlia Perera-Bel, Lara Nonell, Joaquim Bellmunt

## Abstract

Muscle-invasive bladder cancer (MIBC) is associated with poor predictability of response to cisplatin-based neoadjuvant chemotherapy (NAC). Consequently, the benefit of NAC remains unclear for many patients due to the lack of reliable biomarkers. In order to identify biomarkers and build an integrated and accurate model to predict NAC response, we conducted a comprehensive transcriptomic and genomic profiling on 100 MIBC patient tumors.

Our analysis identified 602 differentially expressed genes between responders and non responders. From these, we derived the Top10up and Top10dn gene signatures. We also identified a set of genes related to keratinization (KRT), extracellular matrix (ECM) and cell cycle regulation (CELL CYCLE REG) that were strongly associated with response. Additionally, we found a Wnt-related signature (WNT) significantly associated with non response. Genomic analysis revealed that mutations in DNAH8, DNAH6 or DNAH10 (DNAHalt) and deletions in KDM6A (KDM6Adel) correlated significantly with chemotherapy resistance.

Using our comprehensive molecular analysis as a backbone, we developed different machine learning (ML) models, based on the Xgboost (XGB) approach. The transcriptomics-only model (XGB-R) matched the performance of the combined model (XGB-RW; AUC=0.85). The relative ease of sample collection for transcriptomic data, along with the external validation, make it a promising candidate for clinical translation. To ensure robustness and broader applicability, external validation in larger, more diverse cohorts is necessary. To facilitate this, our most predictive ML models have been made publicly available via GitHub.

**Statement of significance:** We present XGB-R, a robust validated machine-learning model using transcriptomic features to predict neoadjuvant chemotherapy response in muscle-invasive bladder cancer with high accuracy (AUC=0.85), being a candidate for clinical translation.

## INTRODUCTION

Bladder cancer (BC) is a complex disease with a huge impact in society being the 10th most common cancer worldwide and the 6th most common cancer in men. The pathologic tumor stage in localized BC is separated in two main groups according to the invasion into the muscularis propria layer: non-muscle invasive bladder cancer (NMIBC), which includes Tis, Ta and T1, and muscle-invasive bladder cancer (MIBC) tumors, which includes T2, T3 and T4a.

Approximately 20% of newly diagnosed BC patients are MIBC, a clinical setting which is more aggressive than NMIBC and is associated with a poor 5-year survival outcome^3-5^.

Over the last two decades, the standard-of-care for MIBC has been to administer cisplatin-based neoadjuvant chemotherapy (NAC) followed by radical cystectomy (RC). Despite NAC having demonstrated a benefit in overall survival (OS), less than 50% of patients are actually eligible due to comorbidities of frailty, and many of them proceed to RC^6-8^. Moreover, among patients who are treated with NAC, the percentage of non-responders (NR) is around 40%, with these patients having high recurrence rate and poor 5-year survival^3,4^. Despite the great efforts made in recent years, there is still an absolute lack of reliable biomarkers to predict benefit from NAC^4,9^. Consequently, the inability to predict NAC response is a significant barrier to adequately identify those patients who might benefit from it. In turn, this prevents non-responders from avoiding unnecessary toxicity and allows for the provision of alternative treatment options. Identifying and validating integrated and accurate predictive models of NAC is therefore a highly unmet need in MIBC.

BC is associated with a high molecular diversity, and several transcriptomic subtypes have been described^8-13^. The most accepted molecular subtypes were described in the TCGA BC study, in which Robertson et al. proposed five MIBC transcriptomic subtypes (TCGAclas): basal-squamous, luminal papillary, luminal infiltrated, luminal and neuronal^10^. In a parallel study, the Lund taxonomy (LundTax) has been described as an alternative subtype classification: Urothelial-like (Uro), which includes UroA, UroB and UroC, Genomically Unstable (GU), Basal/Squamous (BaSq), Mesenchymal-like (MES-like) and Small-cell/Neuroendocrine (ScNE)^11^. Despite differing in the quantity, proportion and characteristics of the subtypes, both subtype classifications show high overlap^13^ (Supplementary Fig. 1).

Some studies suggest that TCGAclas subtypes predict NAC response, and can be useful for treatment selection and improving outcomes^14^. However, these findings have been demonstrated to be inconsistent over time. First, Choi et al. reported that both basal and luminal were the subtypes showing the greatest benefit from NAC, whereas p53-like tumors were less likely to respond to NAC^9^. On the other hand, Seiler et al. confirmed that basal tumors had the highest benefit of NAC, but conversely indicated that luminal papillary subtype was associated with a low likelihood of benefit from NAC^14^. Additionally, Kamoun et al. highlighted that the luminal infiltrated subtype appeared to be resistant to cisplatin-based therapy but instead particularly sensitive to immune checkpoint inhibitors (ICI)^12^. Moreover, Lotan et al. indicated that non-luminal tumors received the greatest benefit from NAC, while luminal tumors experienced only a minimal survival benefit^15^. Using the LundTax classification, the opposite finding was reported by Sjödahl et al. demonstrating that luminal-like subtypes are the ones that more frequently respond to NAC treatment^16^. This lack of consistent results therefore requires further classification studies to establish the clinical relevance of molecular subtyping in MIBC as predictive biomarkers of NAC^17,18^.

At the genomic level, MIBC is characterized by genomic instability and a high mutation rate^12,18^. The most commonly mutated genes in MIBC are *TP53* (48%), *KMT2D* (28%) and *KDM6A* (26%)^10^. The most common somatic copy number variations (SCNVs) include the amplification of *E2F3*, *PPARG* and *MDM2* and deletions in *CDKN2A* and *RB1^10^*. Importantly, tumor mutational burden (TMB) and APOBEC-mediated mutagenesis have been associated with OS^12^. Mutations in DNA damage response (DDR) genes have been previously associated with response to NAC^19,20^. However, none of these genomic alterations have been validated as potential predictive biomarkers of NAC in MIBC.

In our present study, we have conducted a comprehensive molecular profiling using RNA sequencing (RNA-Seq) and whole exome sequencing (WES) obtained from a cohort of 100 MIBC patients treated with NAC across different hospitals in Catalonia, Spain. Our primary objective was to characterize the transcriptomic and genomic landscape of MIBC in relation to NAC response. Our secondary objective was to develop a machine learning (ML) model capable of predicting the response to NAC treatment. The overarching aim of our study was to create a predictive tool which could be potentially translated into daily practice for selecting ideal candidates for NAC and consequently optimize the management of MIBC patients.

## MATERIALS AND METHODS

### Samples and statistical analyses

Of 130 patients retrospectively collected, only 100 were eligible for molecular profiling (RNA=71, DNA=97). A flowchart of patient selection is provided in Figure 1. FFPE pre-treatment samples were obtained post-TURB. Responders (R) are defined as those achieving <=pT1N0 at cystectomy. Clinical variable comparisons were performed using CompareGroups R package v.4.8.0. Kaplan–Meier survival curves were generated using survival v.3.4.0 and survminer v.0.4.9, with p-values obtained via log-rank test.

**Figure 1.**
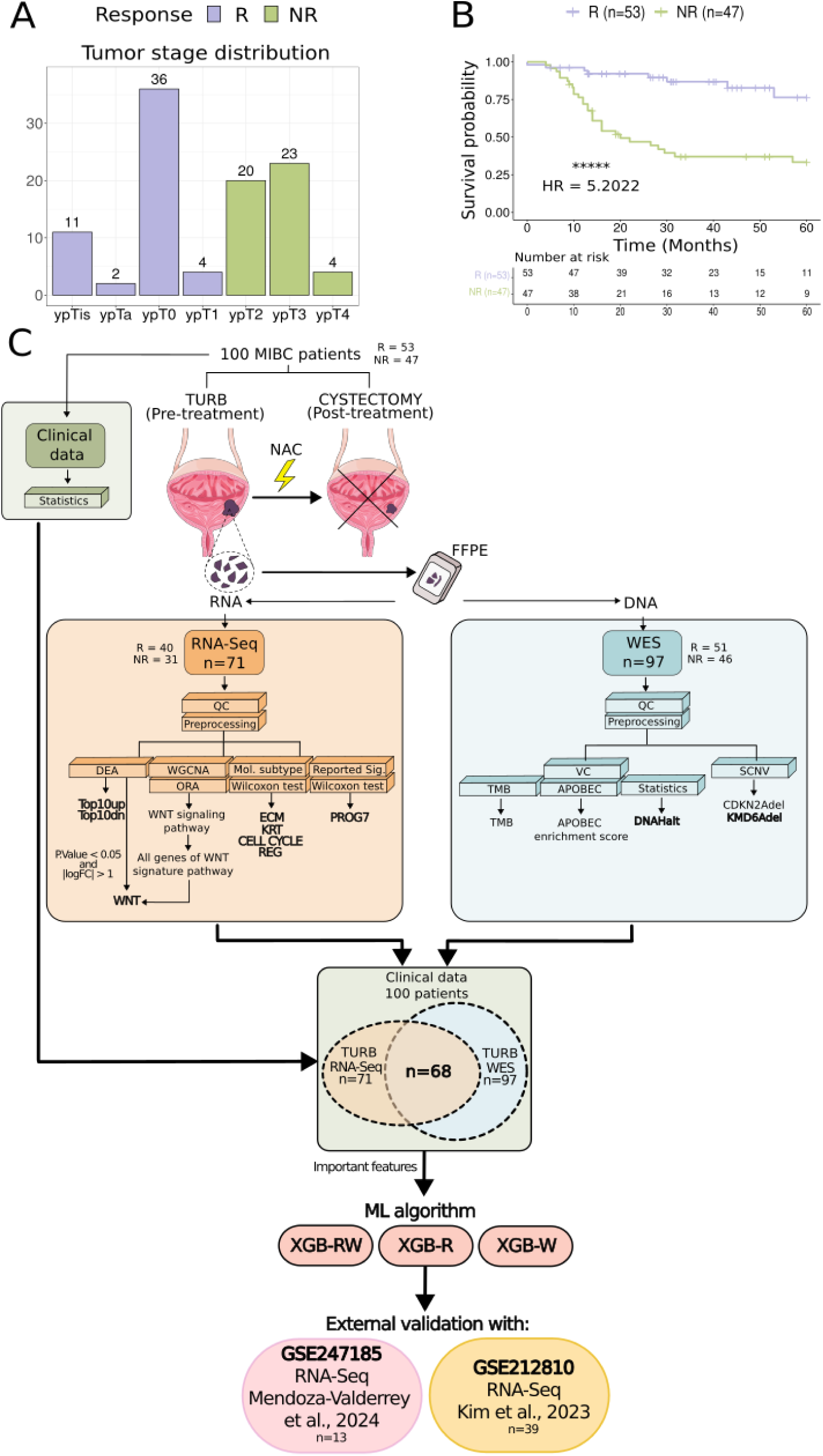
Overview of the MIBC cohort and analysis workflow. **A.** Response and tumor stage distribution across the cohort of 100 MIBC patients. Response was defined as dowstaging to non-MIBC status with no pathological lymph-node involvement (<=T1N0) observed at cystectomy. **B.** Kaplan-Meier survival curve for 5-year overall survival (OS) in MIBC patients stratified by response (p<0.00001; HR=5.20). **C.** Design of the study. A total number of 100 MIBC patients were retrospectively included in the study: 53 responders (R) and 47 non-responders (NR). We generated RNA sequencing (RNA-Seq) (n=71; R=40, NR=31) and whole-exome sequencing (WES) (n=97; R=51, NR=46). Both analyses followed a first quality control (QC) and were subsequently subjected to a preprocessing step. From RNA-Seq data: differential expression analysis (DEA), where two signatures were obtained (Top10up, Top10dn), weighted gene correlation network analysis (WGCNA) followed by an over-representation analysis (ORA) where we obtained a WNT signature associated with NAC response and molecular subtype classification using two classifiers (LundTax and TCGAclas), where two signatures were obtained (ECM, KRT and CELL CYCLE REG). From WES data: variant calling (VC) where we found that mutations in some DNAH family genes (DNAH8, DNAH6 and DNAH10) were significantly associated with response and somatic copy number variation (SCNV) showed that deletion in KMD6A was strongly associated with response. Following the VC, tumor mutational burden (TMB) and APOBEC scores were obtained. Features obtained from RNA-Seq and WES analyses were used to construct three machine learning (ML) models to predict NAC response using data from: 68 patients with both data types for XGB-RW, 71 patients with RNA-Seq data for XGB-R, and 97 patients with WES data for XGB-R. External validation for XGB-R was performed using two external datasets (GSE247185 and GSE212810).

### RNA-Seq Bioinformatics Analysis

Quality control of raw fastq files was performed using fastQC and fastqscreen. STAR v.2.7.8 aligned reads to GRCh38, with annotation from Gencode (hg38 v41). Picard v.2.25.1 assessed alignment quality. Quantification was done with featureCounts (Subread v.2.0.3). Lowly expressed genes (rowSums(counts.m>10) < 42) were filtered. PCA and Hierarchical Clustering were applied to detect and remove outliers.

### Differential Expression Analysis and Weighted Correlation Network analysis (WGCN)

Differential expression was analyzed using Limma v.3.54.2 with TMM normalization (edgeR v.3.40.2). Models included age, sex, and batch as fixed effects; hospital was included as a random effect. Genes were considered differentially expressed at p-value < 0.05 and |logFC| >1. WGCNA (v1.72-5) identified co-expressed gene clusters, being the green one (n=77) significantly correlated to non responders (NR) (Pearson). ORA showed those genes were significantly correlated with the Wnt signaling pathway. Intersection between Wnt-related genes and DE was performed obtaining the WNT signature (n=17)

### Molecular Subtype Classification and RNA-Seq signatures

LundTax and TCGAclas classification was performed using LundTax2023Classifier v.1.1.1 and BLCAsubtyping v.2.1.1. Transcriptomic signature scores obtained through RNA-Seq analysis (Top10up, Top10dn, WNT, ECM, KRT and CELL CYCLE REG) were calculated using GSVA v.1.46.0, with the log2(TPM) table of counts. Then, signatures were compared between R and NR (Wilcoxon test) and performed a survival analysis (survival R package v.3.4.0).

### WES Analysis

Fastq files were processed using nf-core/sarek pipeline v.3.4.0 (default parameters for tumor-only, GRCh38). Quality control was conducted using fastQC v.0.12.1 and fastp v.0.23.4. BWA v.0.7.17 performed alignment, quality-checked with Picard v.2.25.1. Mutect2 (GATK4 v.4.4.0.0) was used for variant calling, followed by filtering with FilterMutectCalls. VCF files were annotated with VEP v.10.2.0 and converted to MAF format using vcf2maf v.1.6.21. Variants were retained if labeled “PASS,” absent from gnomAD v.2.1.1 or with allele frequency (AF) < 0.01, and had a sequencing depth >20.

### Somatic variant calling

MAFtools v.2.18.1 analyzed mutation data. TMB and APOBEC scores were calculated. Significantly differentially mutated genes between R and NR were identified using mafCompare. Survival analysis was conducted based on gene mutation status. Validation of DNAH alterations was performed using cBioPortal.

SCNVs were analyzed using CNVkit v.0.09.10 in tumor-only mode. SCNVs were categorized into five states: 2-copy loss, 1-copy loss, normal, 1-copy gain, and >=2-copy gain. Deletions/amplifications of key genes were statistically compared between R and NR. Survival analysis was performed based on deletions.

### Predictive Models of NAC Response

Different Xgboost (XGB) ML models were constructed (XGB-RW, XGB-W, XGB-R, XGB-WNT), including Top10up, Top10dn, WNT, ECM, KRT and CELL CYCLE REG signatures, DNAH8/6/10 alterations, and KDM6A deletions. The ROC curve represents the mean AUC from 1,000 independent training-test (70%-30%) runs. External validation was performed using GSE237185 (n = 13) and GSE212810 (n = 39) datasets.

## RESULTS

### Patient characteristics and genomic analysis

Our cohort included a total of 100 patients with MIBC retrospectively collected from four Catalan hospitals from 2010 to 2019. All patients received cisplatin-based neoadjuvant chemotherapy (NAC), and were divided into responders (R; n=53) and non-responders (NR; n=47). Response was defined as postsurgical downstaging to non-MIBC (<=ypT1) with no pathological lymph-node involvement (ypN0) observed at cystectomy (Fig. 1A). A detailed summary of clinical and histopathological information is provided in Table1 (and Supplementary Table 1).

Median age of patients (10 females and 90 males) at diagnosis was 69 years (64;75) (Table 1). No differences were observed in the demographic variables between R and NR (Table 1, Supplementary Table 1). Since these patients were treated before the approval of adjuvant nivolumab in Spain^20^, none of them received adjuvant immune checkpoint blockade. There was no difference in the use of additional treatments (prior intravesical therapy, adjuvant chemotherapy) between R and NR (Table 1). The similarity between the administration of adjuvant treatment suggests a consistent approach in the treatment management for R and NR, which is crucial for ensuring unbiased results in further analyses.

**Table 1.** Overview of the clinical characteristics of the MIBC cohort. (Significancy: p<0.05 = *, p<0.01 = **, p<0.001 = ***, p<0.0001 = ****, p<0.00001 = *****)

|  | All patients<br>N=100 | Responders<br>N=53 | Non-responders<br>N=47 | P overall | Sig |
| --- | --- | --- | --- | --- | --- |
| Age (median, range) | 70 [64;75.25] | 69 [64;75] | 70 [64;75] | 0,906 |  |
| Sex (n, %) |  |  |  | 1 |  |
| Male | 90 (90.0%) | 48 (90.6%) | 42 (89.4%) |  |  |
| Female | 10 (10.0%) | 5 (9.43%) | 5 (10.6%) |  |  |
| Prior intravesical treatment (n, %) |  |  |  | 0,465 |  |
| None | 83 (83.0%) | 45 (84.9%) | 37 (78.7%) |  |  |
| Mitomycin | 8 (8.00%) | 3 (5.66%) | 5 (10.6%) |  |  |
| Bacillus Calmette-Guerin | 7 (7.00%) | 4 (7.55%) | 3 (6.38%) |  |  |
| Others | 2 (2.00%) | 0 (0.00%) | 2 (4.26%) |  |  |
| NA | 1 (1.00%) | 1 (1.89%) | 0 (0.00%) |  |  |
| Hydronephrosis at baseline (n, %) |  |  |  | 0,298 |  |
| Yes | 25 (25.0%) | 16 (30.2%) | 9 (19.1%) |  |  |
| No | 75 (75.0%) | 37 (69.8%) | 38 (80.9%) |  |  |
| Number of cycles of neoadjuvant chemotherapy (n, %) |  |  |  | 0,794 |  |
| 1-3 | 83 (83.0%) | 43 (81.1%) | 40 (85.1%) |  |  |
| >=4 | 17 (17.0%) | 10 (18.9%) | 7 (14.9%) |  |  |
| Leukocytes (Median, IQR) | 7435 [5968;8958] | 7800 [5760;8900] | 7410 [6275;8965] | 0,764 |  |
| Neutrophils (Median, IQR) | 4375 [3300;5910] | 4550 [3200;5900] | 4300 [3585;5920] | 0,656 |  |
| Lymphocytes (Median, IQR) | 1700 [1292;2200] | 1710 [1209;2180] | 1600 [1400;2280] | 0,868 |  |
| Ratio neutrophils/lymphocytes (Median, IQR) | 2.42 [1.82;3.77] | 2.44 [1.86;3.14] | 2.40 [1.76;4.29] | 0,882 |  |
| Tumor Stage at TURB (n, %) |  |  |  | 0,811 |  |
| T1 | 5 (5.00%) | 3 (6.38%) | 2 (3.77%) |  |  |
| T2 | 90 (90.0%) | 41 (87.2%) | 49 (92.5%) |  |  |
| T3 | 4 (4.00%) | 2 (4.26%) | 2 (3.77%) |  |  |
| T4 | 1 (1.00%) | 1 (2.13%) | 0 (0.00%) |  |  |
| ypT at cystectomy (n, %) |  |  |  | 0 | **** |
| ypT0 | 36 (36.0%) | 0 (0.00%) | 36 (67.9%) |  |  |
| ypTis | 11 (11.0%) | 0 (0.00%) | 11 (20.8%) |  |  |
| ypTa | 2 (2.00%) | 0 (0.00%) | 2 (3.77%) |  |  |
| ypT1 | 4 (4.00%) | 0 (0.00%) | 4 (7.55%) |  |  |
| ypT2 | 20 (20.0%) | 20 (42.6%) | 0 (0.00%) |  |  |
| ypT3 | 23 (23.0%) | 23 (48.9%) | 0 (0.00%) |  |  |
| ypT4 | 4 (4.00%) | 4 (8.51%) | 0 (0.00%) |  |  |
| ypN Cystectomy (n, %) |  |  |  | 6,48E-10 | **** |
| N0 | 77 (77.0%) | 24 (51.1%) | 53 (100%) |  |  |
| N1 | 7 (7.00%) | 7 (14.9%) | 0 (0.00%) |  |  |
| N2 | 16 (16.0%) | 16 (34.0%) | 0 (0.00%) |  |  |
| Urethral involvement (n, %) |  |  |  | 7,16E-04 | *** |
| Yes | 9 (9.00%) | 0 (0.00%) | 9 (19.1%) |  |  |
| No | 91 (91.0%) | 53 (100%) | 38 (80.9%) |  |  |
| Lymphovascular invasion (n, %) |  |  |  | 9,87E-09 | **** |
| Yes | 24 (24.0%) | 0 (0.00%) | 24 (51.1%) |  |  |
| No | 76 (76.0%) | 53 (100%) | 23 (48.9%) |  |  |
| Surgical margins (n, %) |  |  |  | 0,004 | *** |
| Positive | 7 (7.00%) | 0 (0.00%) | 7 (14.9%) |  |  |
| Negative | 93 (93.0%) | 53 (100%) | 40 (85.1%) |  |  |
| Number of resected lymph nodes at cystectomy (n, %) |  |  |  | 0,307 |  |
| 1-9 | 32 (32.0%) | 16 (30.2%) | 17 (36.2%) |  |  |
| 10-19 | 58 (58.0%) | 34 (64.2%) | 24 (51.1%) |  |  |
| 20-26 | 9 (9.00%) | 3 (5.66%) | 6 (12.8%) |  |  |
| Number of positive lymph nodes at cystectomy (n, %) |  |  |  | 9,87E-09 | **** |
| 1 or more | 24 (24.0%) | 0 (0.00%) | 24 (51.1%) |  |  |
| 0 | 76 (76.0%) | 53 (100%) | 23 (48.9%) |  |  |
| Adjuvant chemotherapy (n, %) |  |  |  | 0,403 |  |
| No | 69 (69.0%) | 39 (73.6%) | 30 (63.8%) |  |  |
| Yes | 31 (31.0%) | 14 (26.4%) | 17 (36.2%) |  |  |
| Local recurrence (n, %) |  |  |  | 0,006 | ** |
| Yes | 11 (11.0%) | 1 (1.89%) | 10 (21.3%) |  |  |
| No | 89 (89.0%) | 52 (98.1%) | 37 (78.7%) |  |  |
| Metastatic recurrence (n, %) |  |  |  | 3,07E-05 | **** |
| Yes | 28 (28.0%) | 5 (9.43%) | 23 (48.9%) |  |  |
| No | 72 (72.0%) | 48 (90.6%) | 24 (51.1%) |  |  |

|  | All patients<br>N=100 | Responders<br>N=53 | Non-responders<br>N=47 | P overall | Sig |
| --- | --- | --- | --- | --- | --- |
| Age (median, range) | 70 [64;75.25] | 69 [64;75] | 70 [64;75] | 0,906 |  |
| Sex (n, %) |  |  |  | 1 |  |
| Male | 90 (90.0%) | 48 (90.6%) | 42 (89.4%) |  |  |
| Female | 10 (10.0%) | 5 (9.43%) | 5 (10.6%) |  |  |
| Prior intravesical treatment (n, %) |  |  |  | 0,465 |  |
| None | 83 (83.0%) | 45 (84.9%) | 37 (78.7%) |  |  |
| Mitomycin | 8 (8.00%) | 3 (5.66%) | 5 (10.6%) |  |  |
| Bacillus Calmette-Guerin | 7 (7.00%) | 4 (7.55%) | 3 (6.38%) |  |  |
| Others | 2 (2.00%) | 0 (0.00%) | 2 (4.26%) |  |  |
| NA | 1 (1.00%) | 1 (1.89%) | 0 (0.00%) |  |  |
| Hydronephrosis at baseline (n, %) |  |  |  | 0,298 |  |
| Yes | 25 (25.0%) | 16 (30.2%) | 9 (19.1%) |  |  |
| No | 75 (75.0%) | 37 (69.8%) | 38 (80.9%) |  |  |
| Number of cycles of neoadjuvant chemotherapy (n, %) |  |  |  | 0,794 |  |
| 1-3 | 83 (83.0%) | 43 (81.1%) | 40 (85.1%) |  |  |
| ≥4 | 17 (17.0%) | 10 (18.9%) | 7 (14.9%) |  |  |
| Leukocytes (Median, IQR) | 7435 [5968;8958] | 7800 [5760;8900] | 7410 [6275;8965] | 0,764 |  |
| Neutrophils (Median, IQR) | 4375 [3300;5910] | 4550 [3200;5900] | 4300 [3585;5920] | 0,656 |  |
| Lymphocytes (Median, IQR) | 1700 [1292;2200] | 1710 [1209;2180] | 1600 [1400;2280] | 0,868 |  |
| Ratio neutrophils/lymphocytes (Median, IQR) | 2.42 [1.82;3.77] | 2.44 [1.86;3.14] | 2.40 [1.76;4.29] | 0,882 |  |
| Tumor Stage at TURB (n, %) |  |  |  | 0,811 |  |
| T1 | 5 (5.00%) | 3 (6.38%) | 2 (3.77%) |  |  |
| T2 | 90 (90.0%) | 41 (87.2%) | 49 (92.5%) |  |  |
| T3 | 4 (4.00%) | 2 (4.26%) | 2 (3.77%) |  |  |
| T4 | 1 (1.00%) | 1 (2.13%) | 0 (0.00%) |  |  |
| ypT at cystectomy (n, %) |  |  |  | 0 | **** |
| ypT0 | 36 (36.0%) | 0 (0.00%) | 36 (67.9%) |  |  |
| ypTis | 11 (11.0%) | 0 (0.00%) | 11 (20.8%) |  |  |
| ypTa | 2 (2.00%) | 0 (0.00%) | 2 (3.77%) |  |  |
| ypT1 | 4 (4.00%) | 0 (0.00%) | 4 (7.55%) |  |  |
| ypT2 | 20 (20.0%) | 20 (42.6%) | 0 (0.00%) |  |  |
| ypT3 | 23 (23.0%) | 23 (48.9%) | 0 (0.00%) |  |  |
| ypT4 | 4 (4.00%) | 4 (8.51%) | 0 (0.00%) |  |  |
| ypN Cystectomy (n, %) |  |  |  | 6,48E-10 | **** |
| N0 | 77 (77.0%) | 24 (51.1%) | 53 (100%) |  |  |
| N1 | 7 (7.00%) | 7 (14.9%) | 0 (0.00%) |  |  |
| N2 | 16 (16.0%) | 16 (34.0%) | 0 (0.00%) |  |  |
| Urethral involvement (n, %) |  |  |  | 7,16E-04 | *** |
| Yes | 9 (9.00%) | 0 (0.00%) | 9 (19.1%) |  |  |
| No | 91 (91.0%) | 53 (100%) | 38 (80.9%) |  |  |
| Lymphovascular invasion (n, %) |  |  |  | 9,87E-09 | **** |
| Yes | 24 (24.0%) | 0 (0.00%) | 24 (51.1%) |  |  |
| No | 76 (76.0%) | 53 (100%) | 23 (48.9%) |  |  |
| Surgical margins (n, %) |  |  |  | 0,004 | *** |
| Positive | 7 (7.00%) | 0 (0.00%) | 7 (14.9%) |  |  |
| Negative | 93 (93.0%) | 53 (100%) | 40 (85.1%) |  |  |
| Number of resected lymph nodes at cystectomy (n, %) |  |  |  | 0,307 |  |
| 1-9 | 32 (32.0%) | 16 (30.2%) | 17 (36.2%) |  |  |
| 10-19 | 58 (58.0%) | 34 (64.2%) | 24 (51.1%) |  |  |
| 20-26 | 9 (9.00%) | 3 (5.66%) | 6 (12.8%) |  |  |
| Number of positive lymph nodes at cystectomy (n, %) |  |  |  | 9,87E-09 | **** |
| 1 or more | 24 (24.0%) | 0 (0.00%) | 24 (51.1%) |  |  |
| 0 | 76 (76.0%) | 53 (100%) | 23 (48.9%) |  |  |
| Adjuvant chemotherapy (n, %) |  |  |  | 0,403 |  |
| No | 69 (69.0%) | 39 (73.6%) | 30 (63.8%) |  |  |
| Yes | 31 (31.0%) | 14 (26.4%) | 17 (36.2%) |  |  |
| Local recurrence (n, %) |  |  |  | 0,006 | ** |
| Yes | 11 (11.0%) | 1 (1.89%) | 10 (21.3%) |  |  |
| No | 89 (89.0%) | 52 (98.1%) | 37 (78.7%) |  |  |
| Metastatic recurrence (n, %) |  |  |  | 3,07E-05 | **** |
| Yes | 28 (28.0%) | 5 (9.43%) | 23 (48.9%) |  |  |
| No | 72 (72.0%) | 48 (90.6%) | 24 (51.1%) |  |  |

The median follow-up was 27 months (IQR 13;112 months); during this time, 1 R and 10 NR patients had local pelvic recurrence, 5 R and 23 NR developed distant metastatic recurrence and 8 R and 30 NR died due to disease. Response groups showed significant differences on 5-year OS (Fig. 1B).

Pathological prognostic features such as urethral involvement, lymphovascular invasion and surgical margins showed significant overrepresentation in NR (p=7.16e-04, p=9.87e-09, p=0.004, respectively) (Table 1). Local and metastatic recurrence were also significantly more common among NR (p=0.006, p=3.07e-05, p=9.87e-09). The ratio between neutrophils and lymphocytes (N/L ratio), previously reported as an indicator of good NAC response^21^, was not significantly different between groups (Table 1).

We generated RNA sequencing (RNA-Seq) data from 71 patients (R=40; NR=31) and whole-exome sequencing (WES) data from 97 (R=51; NR=46) from tumor samples collected at transurethral resection of bladder tumor (TURB) before starting NAC. High-quality sequencing data was obtained for all samples. To ensure the differences in the analyses were not due to initial tumor quality, we confirmed that tumoral fraction and amount of tumor were not statistically different between R and NR. (Supplementary Table 1). RNA-Seq data provided insights into gene expression patterns, molecular subtypes and gene signatures associated with NAC response. The WES data was used to investigate the impact of specific mutations, tumor mutational burden (TMB) and mutational signatures on NAC response. We also studied the significance of somatic copy number variations (SCNVs). Finally, we integrated the information extracted from RNA-Seq and WES data to create a ML model to predict NAC response in MIBC patients (Fig. 1C).

### Wnt gene expression signature is associated to lack of response to NAC

The differential expression analysis between R (n=40) and NR (n=31) identified 602 differentially expressed (DE) genes with a p.value < 0.05 and |logFC| > 1. Among these, a higher number were associated with NR (n=588) compared to a minority that were associated with R (n=14) (Fig. 2A, Supplementary Table 2). To better understand the possible implications for the prediction of response to NAC, and to overcome the lack of adjusted p-values in the previous analysis, we applied the weighted correlation network analysis (WGCNA) method, an unsupervised approach to identify clusters of correlated genes. We identified 14 gene clusters, one of which showed a statistically significant negative association with NAC response (p=0.008, cor=-0.32) (Supplementary Fig. 2A). Genes from this cluster were enriched in several gene sets related to the Wnt signaling pathway, cell signaling, angiogenesis and proliferation (Fig. 2B). Interestingly, we found 17 Wnt signaling pathway genes among the list of DE genes (*SFRP2, FGF10, MIR145, SOX7, ATP6V0C, FOLR1, KREMEN2, TRABD2B, WNT9A, WNT2, AXIN2, PPM1N, SOX2, DLX5, LGR6, LBX2, HESX1*) (Fig. 2B).

**Figure 2.**
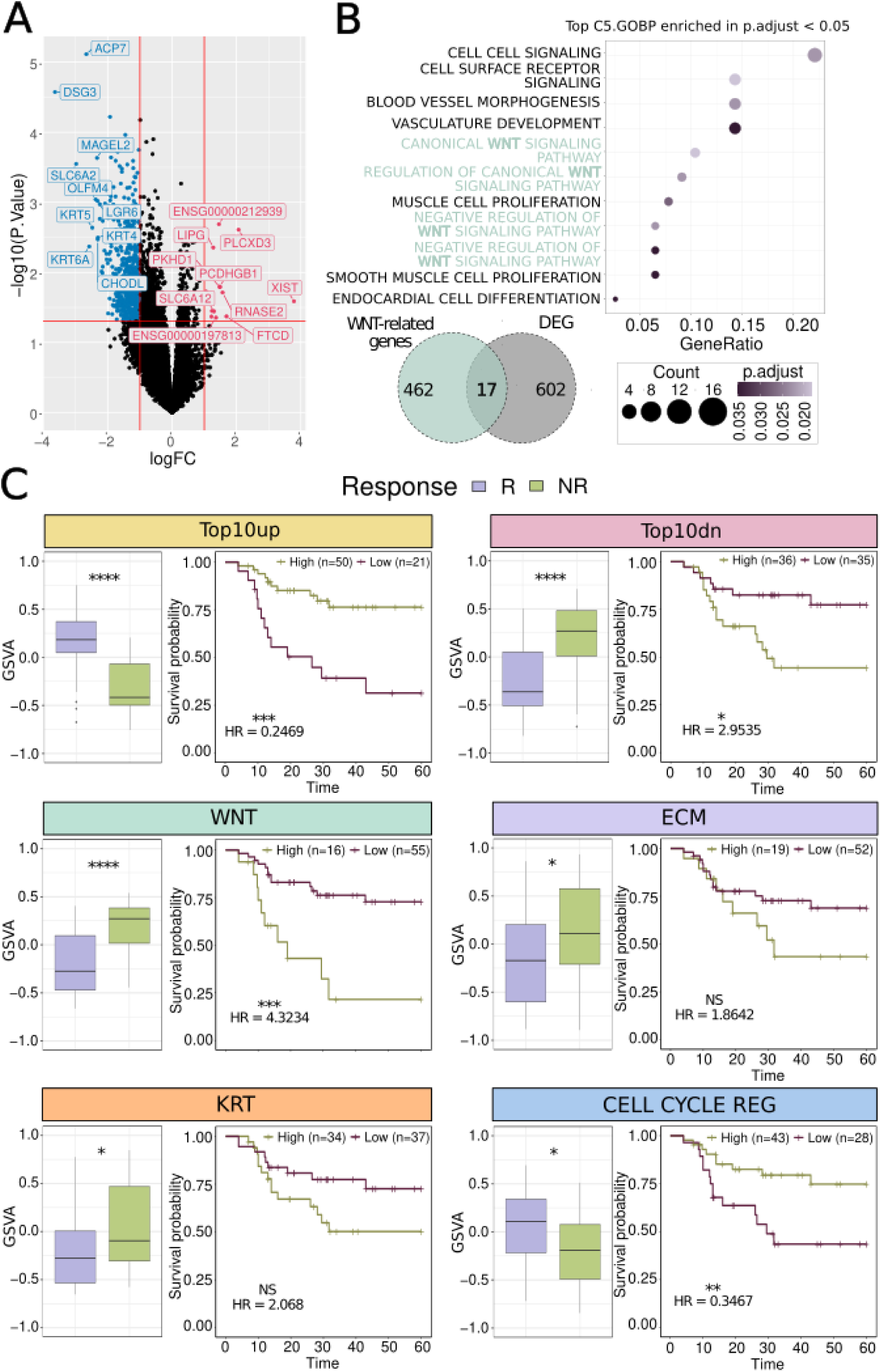
Gene expression signatures are strongly associated with response and 5-year overall survival. **A.** Volcano plot showing the top 10 up-regulated (Top10up) and the top 10 down-regulated (Top10dn) genes obtained after differential expression analysis between responders (R) and non-responders (NR). **B.** Top significantly enriched pathways (p.adj<0.05) in the over-representation analysis (ORA) and 17 genes (WNT signature) resulting from the intersection between DE genes (602) and WNT-related genes (462). **C.** First row: Boxplot of the Top10up (p=1e-07), Top10dn (p=1.2e-05), WNT (p=7.2e-07), ECM (p=0.029) and KRT (p=0.023) and CELL CYCLE REG (p=0.013) signatures score and its significant correlation with NAC response. Second row: Kaplan-Meier curves showing the significant association between Top10up (p=3e-04, HR=0.24[0.10-0.56]), Top10dn (p=0.0129, HR=2.95[1.20-7.22]), WNT (p=2e-04, HR=4.32[1.86-10.02]), ECM ( p>0.05, HR=1.86[0.80-4.31]), KRT (p>0.05, HR=2.06[0.89-4.79]) and CELL CYCLE REG (p=0.0098, HR=0.346[0.14-0.80]) signatures score and 5-year overall survival (OS).

We created signature scores for the top 10 up (Top10up) and down (Top10dn) regulated genes from the DE analysis. As expected, these signature scores showed significant differences between R and NR (p=0.0012,p=0.0002; respectively) (Fig. 2C). Importantly, the gene signature score derived from the 17 Wnt signaling pathway genes also showed a significant difference between R and NR (p=6.2e-07) (Fig. 2C). Beyond the association with NAC response, all three signatures were also capable of stratifying survival outcomes; high expression of the Top10up was associated with better OS (HR=0.38[0.15-0.92]), whereas high expression of Top10dn and WNT signatures were significantly associated with worse OS (HR=3.24[1.42-7.69] and HR=4.53[1.92-11.1] (respectively)(Fig. 2C).

### Molecular subtype signatures provide further insights into response to NAC

We applied the molecular subtype classification from Robertson et al., 2017. TCGAclas classified the samples as Basal squamous (n=18; R=11, NR=7), Luminal (n=12; R=4, NR=8), Luminal infiltrated (n=13; R=6, NR=7) and Luminal papillary (n=28; R=19, NR=9). Although no significant differences were found between R and NR patients, we observed a trend towards a better response to NAC in luminal papillary and basal squamous subtype (68% and 61% of responders, respectively), compared to luminal (33% responders) and luminal infiltrated (46% responders) (Supplementary Fig. 3A, Supplementary Table 3).

We also applied the molecular subtype classification from LundTax and identified the largest group of samples belonging to the Uro group (n=44, 62%); including the different subgroups of: UroA (n=28), UroB (n=7) and UroC (n=9). 13 (18.3%) were classified as BaSq, 12 (16.9%) as GU and 2 as ScNE (2.8%) (Supplementary Fig. 3B). These proportions are in line with the original classification.

None of LundTax subtypes showed a significant association with response (Supplementary Table 3), but proportions were in line with what has been described in the literature: patients classified as UroB (3 R vs 4 NR) and BaSq (5 R vs 8 NR) showed a low proportion of responders (approximately 40%), indicating a lower likelihood of responding to NAC. On the contrary, GU (8 R vs 4 NR) and UroA (17 R vs 11 NR) patients showed an opposite trend, with more than 60% of responders being the 2 subtypes most likely to benefit from NAC (Supplementary Fig. 3B). In addition, whereas luminal papillary in TCGAclas is consistent with the findings in the UroA LundTax subtype, BaSq results contradict our previous findings. This is likely because the BaSq TCGAclas subtype contains patients classified as GU in LundTax, which may increase the proportion of responders in this TCGAclas subtype (Supplementary Fig. 3C).

To understand the response mechanisms within the molecular subtypes, we derived single sample scores of the individual signatures used by the LundTax and TCGA classifiers (Supplementary Fig 2B, 3B). Interestingly, signatures of extracellular matrix (ECM), keratinization process and cell cycle regulation (ECM including PGM5, DES, C7, SFRP4, COMP, SGCD genes^10^, KRT including VSNL1, KRT14, TGM1, SERPINB4, GSDMC, KRT6A, LGALS7, SFN, SPRR2A, BG205162, C12orf54, SPRR2D, HOXD11, KRT6C, KRT5, DSG3, KRT6B, HOXD10, IL20RB, RHCG, AHNAK2, SPRR2F, FGFBP1 genes^13^ and CELL CYCLE REG including FGFR3, CCND1, E2F3, RB1 and CDKN2A genes^13^) showed the most highly significant differences between R and NR, being positively associated with response and OS. The association between ECM and KRT signatures and OS was not significant (Fig. 2C).

### The combination of gene expression signatures stratifies response and overall survival

To assess the potential of these signatures as biomarkers for a predictive model, we studied the correlation between the genes of all the identified signatures. High correlation coefficients were seen in three genes from the Top10dn signature (KRT6A-KRT5=0.75, KRT6B-KRT6C=0.80, SFRP4-SFRP2=0.83 Fig. 3A). KRT6A, KRT6B, KRT6C and KRT5 belong to the keratin family of proteins, which are essential markers for basal epithelial cells. Moreover, most of the keratin family genes were correlated among them (∼-0.60) (Fig. 3A). Additionally, SFRP4 and SFRP2, from the ECM signature, are both part of the secreted frizzled-related protein family. Besides that, there was an overall weak association among the genes from the identified signatures (|cor| <= 0.65, Fig. 3A, Supplementary Table 4), suggesting a need to further explore their potential as combined biomarkers.

**Figure 3.**
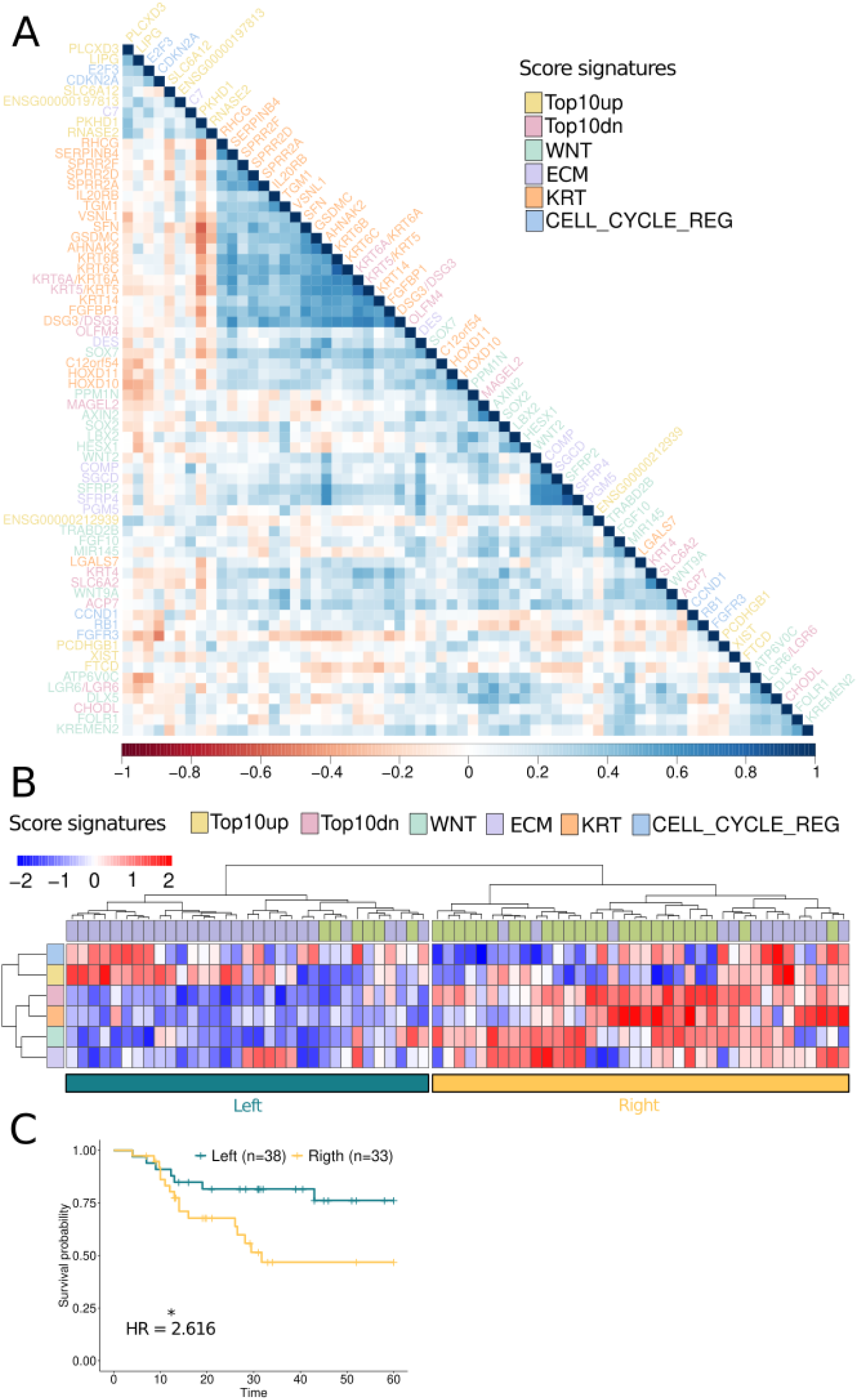
Gene expression signatures can distinguish between R and NR. **A.** Correlation plot of the expression of genes constituting the RNA-Seq signatures. **B.** Heatmap showing the scores of RNA-Seq signatures across samples and their impact in response. **C.** Kaplan-Meier showing the significant association between clusters obtained in heatmap and 5-year overall survival (OS) (p=0.0287, HR=2.616[1.07-6.39]).

The combination of the six signatures by unsupervised hierarchical clustering identified two main clusters of patients (Fig. 3B). The left cluster (Fig. 3C), which mainly contained R, was characterized by low expression of WNT, Top10dn, KRT and CELL CYCLE REG signatures and overexpression of ECM and Top10up signatures. The right cluster (Fig. 3C) contained the majority of NR and was characterized by an overexpression of all the signatures except the CELL CYCLE REG and KRT. Within the later cluster, we could further distinguish a subcluster with high expression of CELL CYCLE REG signatures that could discriminate against the few responders with high WNT signature values. In line with the gene-wise correlation results, we observed that the signatures captured distinct relevant aspects associated with NAC response, harboring strong potential features for a predictive model. Similarly to the individual signatures, the combination of the six signatures led to the differentiation of two groups with significantly different 5-year OS (p=0.0287, HR=2.616[1.07-6.39]) (Fig. 3C).

### Analysis of mutational landscape

Analysis of WES data identified a total of 27.404 somatic non-synonymous mutations in our cohort. The most frequently mutated genes were KIR2DL3 (54%) and LILRB3 (54%), followed by TP53 (51%), TTN (48%), RAMEF18 (46%), CYP2D6 (44%), MUC3A (41%), MUC5AC (41%), GTPBP6 (38%) and ZBED3 (38%) (Supplementary Fig. 4A).

The most frequent somatic mutations in 121 previously reported BC associated genes (SMG) (Supplementary Table 5) were TP53, with 49 mutated patients (51%), followed by KMT2D (24%), ERCC2 (18%), EP300 (16%) and ATM (15%) (Fig. 4A). This frequency is consistent with previously reported SMG in other MIBC cohorts^10,21-23^.

**Figure 4:**
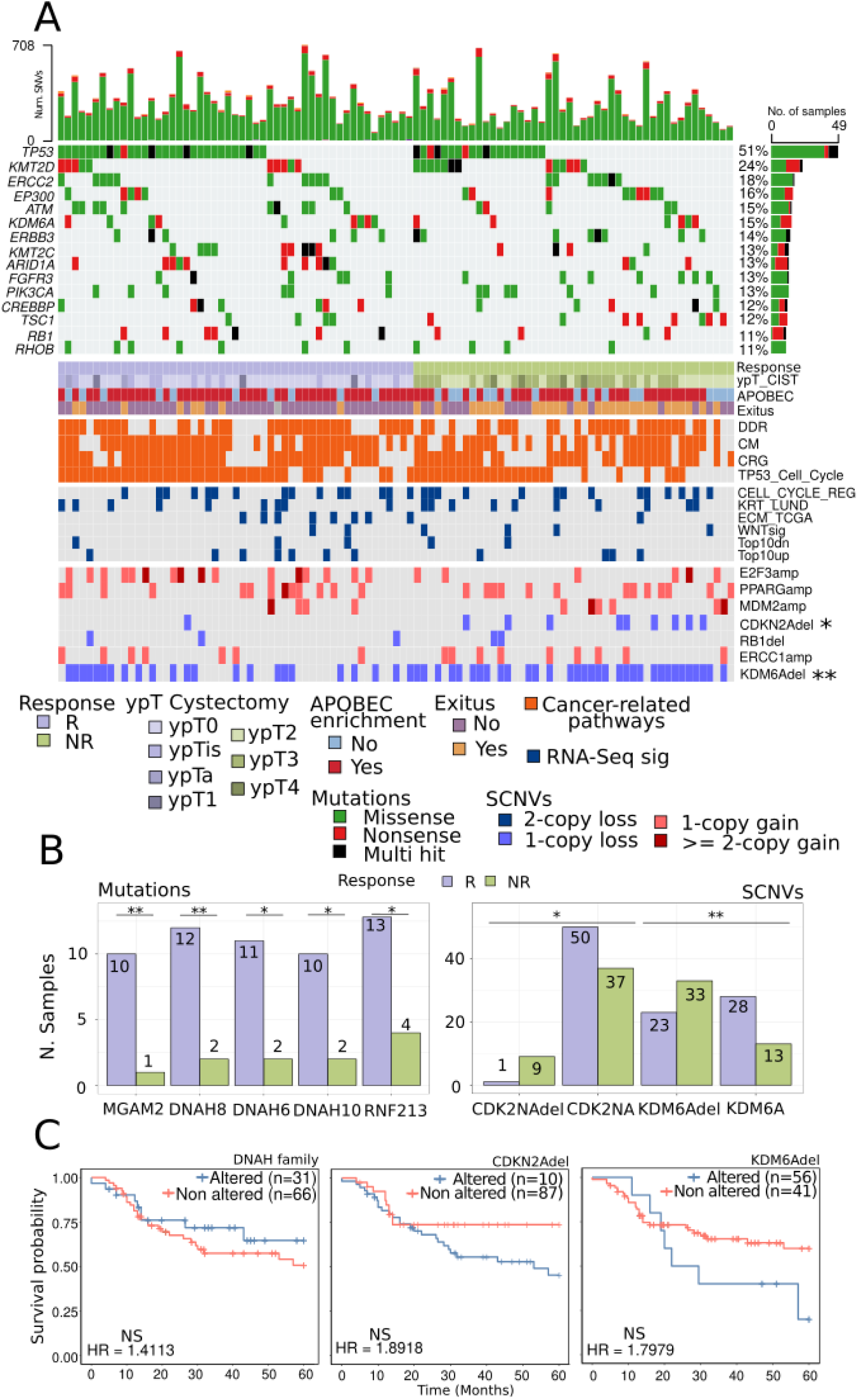
Mutations in DNAH family genes (DNAH8, DNAH6 and DNAH10) and deletions in CDK2NA and KDM6A are significantly correlated with NAC response. **A.** Mutational landscape for MIBC patients with clinical information. Bladder cancer genes mutated in more than 10% of samples. Mutations in important pathways such as DDR, CM, CRG and TP53CellCycle as well as RNA-Seq signatures were also included. Somatic copy number variations (SCNVs) previously identified as important features were also visualized in the oncoplot. **B.** Boxplots of significantly different mutated genes associated with NAC response: MGAM2 (p=0.0086), DNAH8 (p=0.0086), DNAH6 (p=0.0163, DNAH10 (p=0.0301), RNF213 (p=0.0349) and deletions in CDK2NA (p=0.006) and KDM6A (p=0.014) and its significant association with NAC response. **C.** Kaplan-Meier curve showing the correlation of: mutations in any of the DNAH family genes (DNAH8, DNAH6 and DNAH10) (p>0.05) and deletions in CDKN2A (p>0.05) and KDM6A (p>0.05) with 5-year overall-survival.

Cancer pathways that have been associated with BC tumorigenesis (e.g. DDR, CM, CRG, TP53) (Supplementary Table 6) were mutated in the majority of patients in our cohort. In contrast, the signatures obtained from RNA-Seq analysis (top10up, top10dn, WNT, KRT, ECM and CELL CYCLE REG) were mutated in a small proportion (10-20%) of patients. Overall, the mutational landscape at the pathway level remained similar across the entire cohort and it was independent of response (Fig. 4A).

### Somatic mutations in DNAH8, DNAH6 and DNAH10 genes are correlated with response to NAC and OS

We tested for association between the mutational profile and response to NAC (R n=51, NR n=46). None of the frequently mutated genes nor pathways showed significantly different proportions between R and NR. However, we found significant differences in five genes not previously reported to be related to response to NAC in MIBC (Fig. 4B, Supplementary Table 7, Supplementary Fig. 4B,5A): MGAM2 (R=10, NR=1; p=0.0086), DNAH8 (R=12, NR=2; p=0.0086), DNAH6 (R=11, NR=2; p=0.0163, DNAH10(R=10, NR=2; p=0.0301), RNF213 (R=13, NR=4; p=0.0349) (Supplementary Fig. 5A). As expected, mutations in DNAH family genes (DNAH8, DNAH6 and DNAH10) tend to co-occur, while mutations in RNF213 and DNAH genes are mutually exclusive. Moreover, mutations in MGAM2 and RNF213 have a high co-occurrence (Supplementary Fig. 5B). Survival analysis of these 5 genes did not yield statistically significant results (Supplementary Fig. 6A), probably due to the low frequency of these events. Grouping patients with mutations in any of DNAH8, DNAH6 and DNAH10 (DNAHalt) did not reach statistical significance in our cohort either (p=0.369; HR=1.41[0.60-3.00], Fig. 4C). To overcome the limitation due to the low frequency of the events and further investigate the potential prognostic value of these genes, we used different publicly available BC datasets from cBioPortal. DNAHalt showed a positive association with better 5-year OS (p=3.89e-04; HR=1.62[:1.29-2.02]) (Supplementary Fig. 6B). Those results suggest that somatic mutations in any of the DNAH8, DNAH6 or DNAH10 genes might predict not only NAC sensitivity, but also better OS.

Biomarkers of response to platinum-based chemotherapy reported in other clinical contexts, such as in the metastatic settings like tumor mutational burden (TMB, total mutations per Mb) and APOBEC mediated-mutation scores (APOBECscore) could not effectively distinguish between R and NR in our cohort. APOBECscore did not show a significant correlation with 5-year OS either, whereas TMB was significantly associated with 5-year OS (p=0.0231, HR=2.1636[1.09-4.28], suggesting that TMB has a prognostic rather than a predictive value (Supplementary Fig. 6C).

### Deletions in CDKN2A and KDM6A are correlated with response

SCNVs play a crucial role in cancer development, progression and treatment resistance/sensitivity. Amplifications and deletions in genes frequently implicated in BC (E2F3amp, PPARGamp, MDM2amp, ERBB2amp, CDKN2Adel and RB1del)^10,21-23^ occurred across the whole cohort. ERCC1amp and KDM6Adel have previously been identified for their significant role in response to platinum-based chemotherapy^24,25^. Among the SCNVs studied, only CDKN2Adel and KDM6Adel were statistically significantly more prevalent in NR (p=0.014) (Fig. 4B, Supplementary Table 8). However, survival analysis did not show any significant association between these SCNVs and 5-year OS (Fig. 4C). These findings suggest deletions in CDKN2A and KDM6A may potentially serve as predictive biomarkers for NAC response rather than acting as prognostic markers.

### Molecular features can predict NAC response in MIBC

Gene expression signatures from RNA-Seq analysis (Top10up, Top10dn, WNT, ECM, KRT, CELL CYCLE REG) and genomic events from WES (mutations in any of DNAH8, DNAH6 or DNAH10 and deletions in KDM6A) were used to construct a machine learning (ML) algorithm to predict response to NAC (Supplementary Table 9). The XGBoost (XGB) model trained with both RNA and WES features (XGB-RW) achieved an area under the curve (AUC) of 0.85 (Fig. 5A, Supplementary Table 10). Importantly, XGB models offer insights into feature importance, which indicates how much every feature contributes to predicting the target variable. The most contributing features were Top10up (0.282) and WNT (0.203) signatures, followed by Top10dn (0.122) and ECM (0.121) signatures (Fig. 5B, Supplementary Table 10). The variables that contributed less to the model were KDM6Adel (0.104) and CELL CYCLE REG (0.067), followed by DNAHalt (0.064) and KRT (0.04) (Fig. 5B, Supplementary Table 10). Based on that, we tried three different models, a model with only WNT signature (XGB-WNT) (AUC=0.74, Fig. 5A, Supplementary Table 10), a model with only RNA-Seq variables (XGB-R) (AUC=0.85, Fig. 5A, Supplementary Table 10) and a model with only WES features (mutations and SCNVs) (XGB-W) (AUC=0.72, Fig. 5A, Supplementary Table 10).

**Figure 5:**
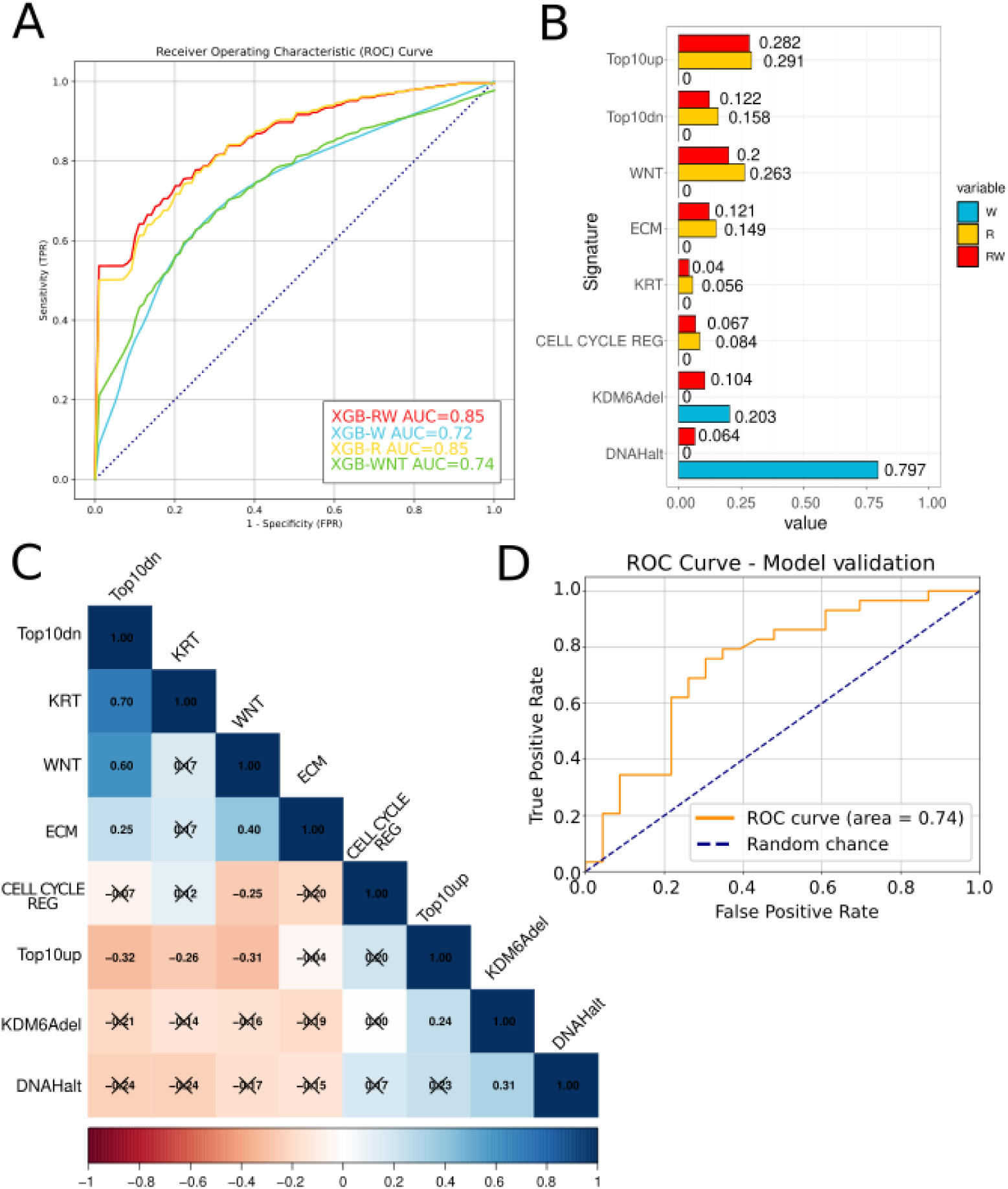
Machine learning models with RNA-Seq and WES data to predict response to NAC. **A.** Synthetic Receiver Operating Curves (ROC) of four Xgboost (XGB) models combining RNA-Seq and WES data (XGB-RW, XGB-R, XGB-W and XGB-WNT). The respective AUCs are: 0.85, 0.85, 0.72 and 0.74. **B.** XGB-RW, XGB-R and XGB-W features ordered by decreasing feature importance. **C.** Correlation analysis of features from RNA-Seq and WES data included in the XGB models (Significancy: p>0.05 X). **D.** External validation of the XGB-R model using GSE237185 (n=13) and GSE212810 datasets (n=39).

We interrogated the correlation among the model variables to better understand the feature importance and to assess whether we were dealing with redundant features. We observed an overall weak association among the features (Fig. 5C), which would support the inclusion of all features in the model as they capture distinct phenotypes. However, an exception was noted for the KRT and WNT signatures, which showed a strong negative correlation with the Top10dn signature (|cor| = 0.7 and 0.6) (Fig. 5C).

Notably, two publicly available datasets from the Gene Expression Omnibus (GEO) database (GSE237185, n=13; GSE212810, n=39) were used to independently validate the XGB-R model. The model achieved an AUC of 0.74 on the combined dataset (Fig. 5D), and AUCs of 0.71 and 0.75 when evaluated separately on each dataset, respectively (Supplementary Fig. 6D).

While some studies have shown that molecular subtypes could be potentially used to personalize BC treatment^14,17^, we achieved poor AUCs for XGB models using the LundTax and TCGAclas subtypes (0.47 and 0.56, respectively; Supplementary Fig. 6E, Supplementary Table 10), suggesting that molecular subtypes alone were unable to accurately predict response to NAC.

## DISCUSSION

Identifying robust and accurate predictive biomarkers for neoadjuvant chemotherapy (NAC) response remains a critical unmet need in muscle-invasive bladder cancer (MIBC). Our study analyzed transcriptomic and genomic profiles of 100 MIBC patients treated with cisplatin-based NAC, correlating molecular features with treatment response (≤pT1N0). This is among the largest real-world studies investigating transcriptomic and genomic determinants of NAC benefit in MIBC.

A key finding is the identification of a novel gene signature involving the Wnt signaling pathway, strongly associated with non-response to NAC. Dysfunction in Wnt/β-catenin signaling is implicated in tumor progression and chemoresistance^26-28^. Our results highlight the WNT signature as a promising biomarker for predicting NAC response, demonstrating high predictive power. Importantly, this finding reinforces the potential of Wnt inhibition as a therapeutic approach in MIBC. While no clinical trials targeting Wnt are available in bladder cancer disease, studies in other cancers suggest its potential in overcoming chemotherapy resistance. The Wnt inhibitor PRI-724 has been tested in pancreatic, colorectal, and myeloid cancers, though concerns remain regarding adverse effects and patient selection^29^. Further investigation will be needed after optimizing dosing and to evaluate the clinical utility of Wnt inhibitors in MIBC.

Most of the prior studies in MIBC have been focused on finding potential biomarkers, molecular subtypes or signatures which can be associated with response to cisplatin-based NAC in MIBC^10,12,14,16,30-32^. However, only few models are publicly available to predict cisplatin-based NAC response using transcriptomics, and although molecular subtypes derived from transcriptomic profiling have been proposed, attempts to correlate them with treatment response have yielded inconsistent results^10,12,14-17^. Among our predictive models, the XGB-R model emerged as the most robust, achieving the highest AUC (0.85) and being validated by two independent public datasets. This model based on transcriptomic features achieves predictive accuracy similar to that of XGB-RW, while surpassing previously reported classifiers^31,32^. The importance of transcriptomic variables, particularly the Top10up, and WNT signatures, suggests that molecular profiling can enhance clinical decision-making in NAC treatment. Given the promising predictive performance of XGB-R, this model, upon external validation, holds potential for clinical translation, providing oncologists with a practical tool for stratifying patients based on NAC benefit.

Based on the findings on CELL CYCLE REG and Top10up signatures, our results support the hypothesis that highly proliferative tumors are more vulnerable to NAC. Several upregulated genes in responders, including long non-coding XIST, LIPG and SLC family members, are associated with proliferation, metabolism, and tumor growth, though not all are directly linked to bladder cancer^33-35^. Conversely, genes such as RNASE2, FTCD, PKHD1, and PCDHGB1, also upregulated, are linked to tumor suppression and favorable prognosis^36-39^. The observation that half of the top 10 upregulated genes in R are associated with proliferation and tumor growth, while the other half are tumor suppressor genes linked to improved prognosis, suggests a dual regulatory pattern. While upregulation of proliferation-associated genes may reflect a high intrinsic tumor cell proliferation rate, which is known to increase sensitivity to cytotoxic chemotherapy^40^, the upregulation of tumor suppressor genes could reflect that tumors may have an intrinsic vulnerability, making them more prone to chemotherapy-induced cell death^41^.

Most of the top 10 downregulated genes in R are linked to basal-squamous differentiation and keratinization. Key markers such as DSG3, KRT4, KRT5, and KRT6A suggest a strong basal-like phenotype and aggressiveness in NR tumors^41-45^, aligning with prior studies that indicate limited therapeutic benefit from NAC in such subtypes^16^. A high keratinization (KRT) signature in NR further supports its role in treatment resistance^46^. Notably, NR tumors also show elevated expression of extracellular matrix (ECM) genes, which are linked to enhanced survival, invasion, and chemoresistance^47^, reinforcing the potential of basal, keratinization, and ECM markers as predictors of poor response to NAC in bladder cancer.

The overall mutational landscape of our cohort was consistent with those already reported in prior studies^10,21-23,48^. Globally, mutations in DDR genes known to correlate with response to cisplatin-based NAC^19^ in prior studies, yet did not emerge as relevant predictors of NAC response in our study^30,49^. ERCC2, RB1 and ATM have been used to enrich platinum responders in bladder preservation studies^30,19,50,51^. In our cohort, these three genes were more frequently mutated in R than NR but without statistical significance. These findings are consistent with some studies in the literature^30,49-51^. Tumor heterogeneity, together with other factors such as epigenetic modifications, immune response or tumor microenvironment, might also influence the NAC response, difficulting the detection of clear mutation-response associations as a single factor^10,12,49,50^.

We identified novel mutations in five genes (MGAM2, DNAH8, DNAH6, DNAH10, and RNF213) with potential relevance to NAC improved response. DNAH gene mutations, involved in cell motility, have already been correlated with chemotherapy sensitivity in gastric cancer^52^, while MGAM2 has been implicated in immune response modulation^53^. The functional role of these genes and their impact in cisplatin sensitivity will require future studies. Somatic copy number variation (SCNV) analysis revealed significant associations with NAC resistance. Specifically, KDM6A deletions correlated with non-response, consistent with its role in DNA damage repair and chemotherapy resistance^25,54^. Similarly, CDKN2A loss was significantly linked to NAC resistance, aligning with prior studies demonstrating its role in tumor progression and, potentially, immunotherapy resistance^55^. While the relationship between CDKN2A deletion and chemotherapy response is less well-established, our findings warrant further investigation into its potential negative predictive value.

Although our study presents compelling findings, certain limitations must be acknowledged. The relatively small sample size increases the risk of overfitting and may limit generalizability. Additionally, the absence of matched normal tissues complicates the distinction between somatic and germline mutations. Despite implementing rigorous filtering strategies, some false-positive variant calls may persist. Moreover, response definitions across studies vary, introducing additional heterogeneity that may impact reproducibility. To address these challenges, we emphasize the need for prospective validation in larger and more diverse cohorts. The robust internal validation by bootstrap .364+ of our models, and particularly the external validation of XGB-R, supports their reliability. Yet external validation in larger datasets for the four models is crucial for clinical implementation. Future studies should explore the integration of our predictive models with other diagnostic tools, including imaging and liquid biopsy markers, to enhance accuracy and real-world applicability.

In conclusion, the XGB-R model, with its high predictive accuracy, comprehensive data integration and external validation, represents a promising candidate for translation into routine oncology practice. Its implementation could enable personalized NAC strategies, improving treatment selection and minimizing unnecessary toxicity for non-responders. Additionally, our results highlight the potential role of Wnt inhibition as an alternative treatment for chemotherapy-resistant MIBC patients.

Future efforts will concentrate on translating this predictor into clinical practice, validating its effectiveness in real-world healthcare settings and incorporating it into routine patient care workflows. By leveraging transcriptomic profiling, we can move closer to a precision medicine approach, optimizing NAC strategies to improve patient outcomes in MIBC.

## Data availability

We have made available four independent ML models (XGB-RW, XGB-R, XGB-W and XGB-WNT), accessible via our GitHub repository (ongoing). RNA-Seq and WES data are available in EGA (ongoing).

## Data Availability

All data produced in the present study are available upon reasonable request to the authors

## Acknowledgements

The work was supported by the following grants and agencies: Projects PI19/00004 and PI22/00171, funded by Instituto de Salud Carlos III (ISCIII) and co-funded by the European Union; a grant from FIS-ISCIII (FI20/00095), 2021SGR00042 by Generalitat de Catalunya;

This work was also supported by the “Xarxa de Bancs de Tumors” sponsored by Pla Director d’Oncologia de Catalunya (XBTC). We want to particularly acknowledge the patients and the IGTP-HUGTP Biobank integrated in the Spanish National Biobanks and BiomodelsNetwork of Instituto de Salud Carlos III (PT20/00050) and Tumor Bank Network of Catalonia for its collaboration.

## Competing interests

Potential conflicts of interest: J. Bellmunt has served in consulting or advisory roles for Astellas Pharma, AstraZeneca/MedImmune, Bristol Myers Squibb, Genentech, Novartis, Pfizer, Pierre Fabre, and the healthcare business of Merck KGaA, Darmstadt, Germany; has received travel and accommodation expenses from Ipsen, Merck & Co., Kenilworth, NJ, and Pfizer; reports patents, royalties, other intellectual property from UpToDate; reports stock and other ownership interests in Rainier Therapeutics; has received honoraria from UpToDate; and has received institutional research funding from Millennium, Pfizer, Sanofi, and the healthcare business of Merck KGaA, Darmstadt, Germany. A. Rodriguez-Vida has served in consulting or advisory roles for Astellas Pharma, Bristol Myers Squibb, Novartis, Pfizer, Johnson&Johnson, Merck, Bayer, MSD, Ipsen; has received travel and accommodation expenses from Ipsen, Merck, Johnson&Johnson and Bayer.

## Notes

### Funding Statement

The work was supported by Project PI19/00004 and grant FI20/00095, funded by Instituto de Salud Carlos III (ISCIII) and co-funded by the European Union; 2021SGR00042 by Generalitat de Catalunya

### Author Declarations

Comite Etica de la Investigacio de l Hospital Universitari Germans Trias i Pujol Comite Etico de Investigacion Clinica del Hospital del Mar

### Summary of Updates

This version of the manuscript has been revised to incorporate substantial improvements that enhance the robustness and relevance of the findings. Key revisions include a change in the methodology used to calculate gene set scores, which now allows for more accurate and biologically meaningful interpretation of the data. Additionally, we have updated the study with new gene signatures that are more reflective of the current understanding of the underlying biological processes, leading to improved sensitivity and specificity in our analyses. Finally, we have implemented an external validation strategy using independent datasets to confirm the reproducibility and generalizability of our results. Together, these changes significantly strengthen the overall impact and reliability of the manuscript.

