## Supplementary material for "Transcriptomics-Driven Machine Learning Models Accurately Predict Chemotherapy Response in Muscle-invasive Bladder Cancer": Supplementary_Figures.pdf

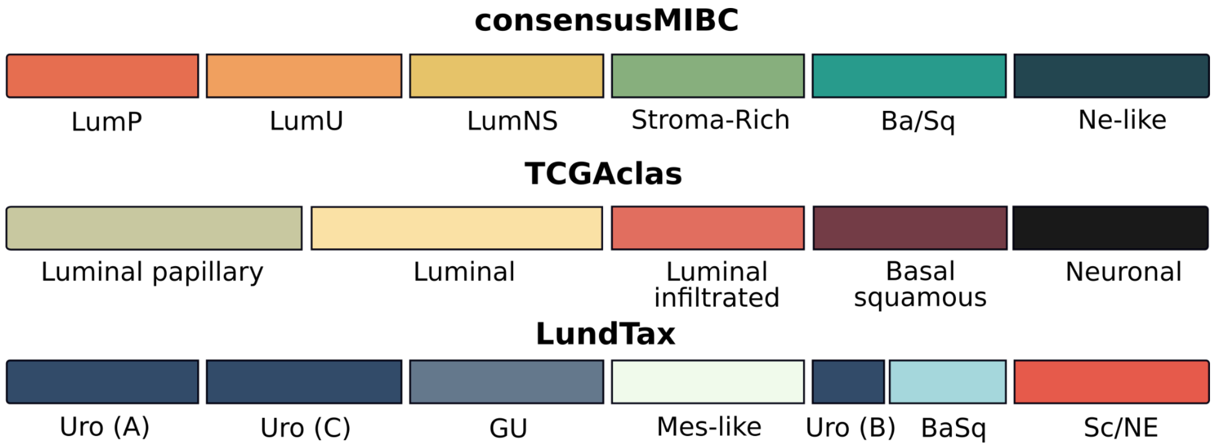


***Supplementary Figure 1. Concordance of bladder cancer molecular subtypes across three different classifiers.*** *The scheme shows the distribution and overlap of molecular subtypes in muscle-invasive bladder cancer as defined by the ConsensusMIBC, TCGAclas, and LundTax classifiers. Subtype assignments are shown for each classifier, highlighting areas of concordance and divergence across the three methods.*

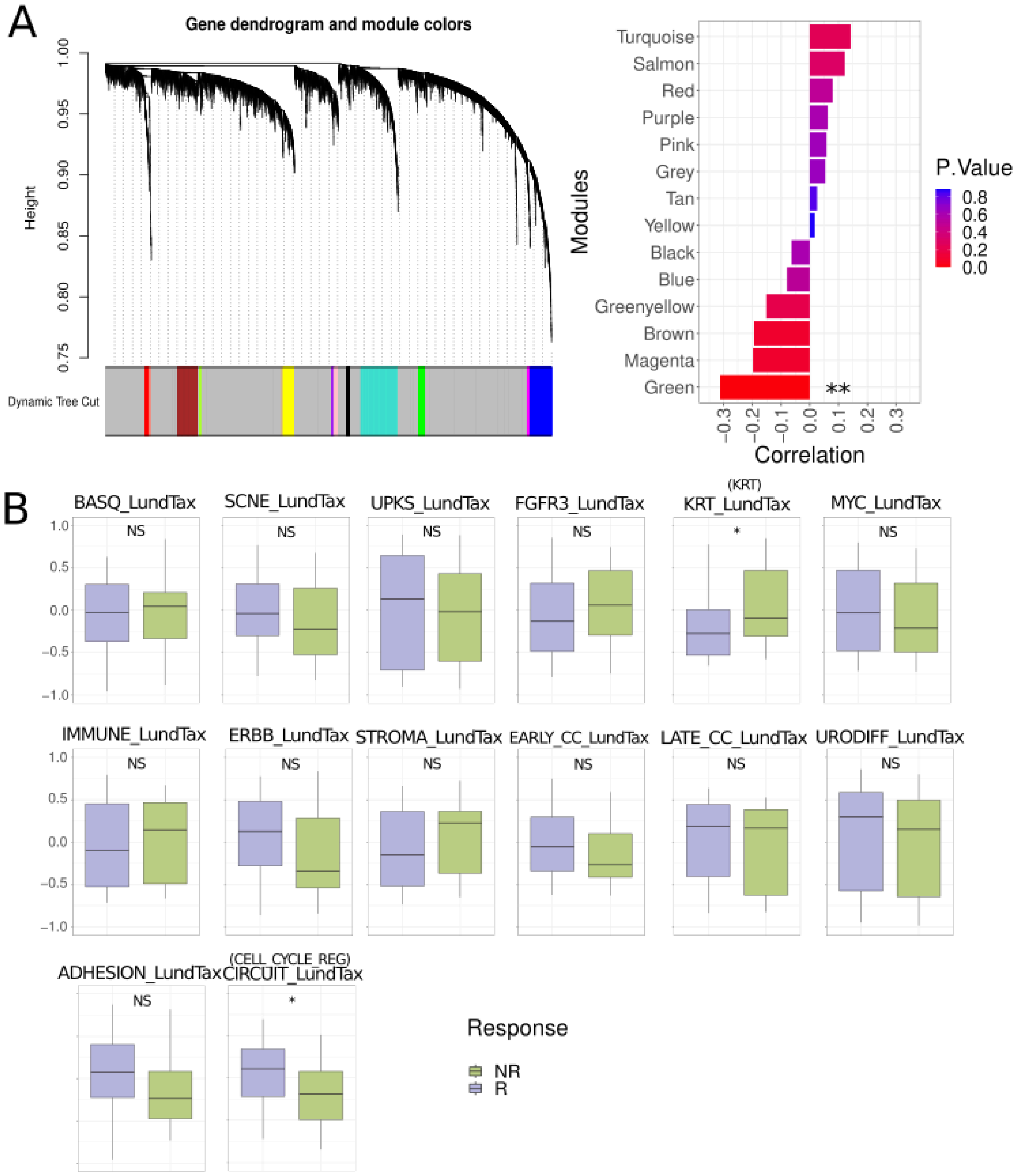


***Supplementary Figure 2. WGCNA results and LundTax classifier signatures. A.*** *Dendrogram showing the modules obtained by weighted gene correlation network analysis (WGCNA) (n=14) and barplot showing the significant negative correlation between green module and response* (p=0.008, cor=-0.32*). **B.** Boxplots showing the differences between R and NR in scores obtained by GSVA for different signatures that LundTax uses for molecular subtype classification (BASQ_LundTax, SCNE_LUND, UPKs_LundTax, FGFR3_LundTax, MYC_LundTax, IMMUNE_LundTax, ERBB_LundTax, STROMAL_LundTax, EARLY_CC_LundTax, Late_CC_LundTax, URODIFF_LundTax, ADHESION_LundTax, p>0.05. KRT_LundTax p=0.023, CIRCUIT_LundTax p=0.013).*

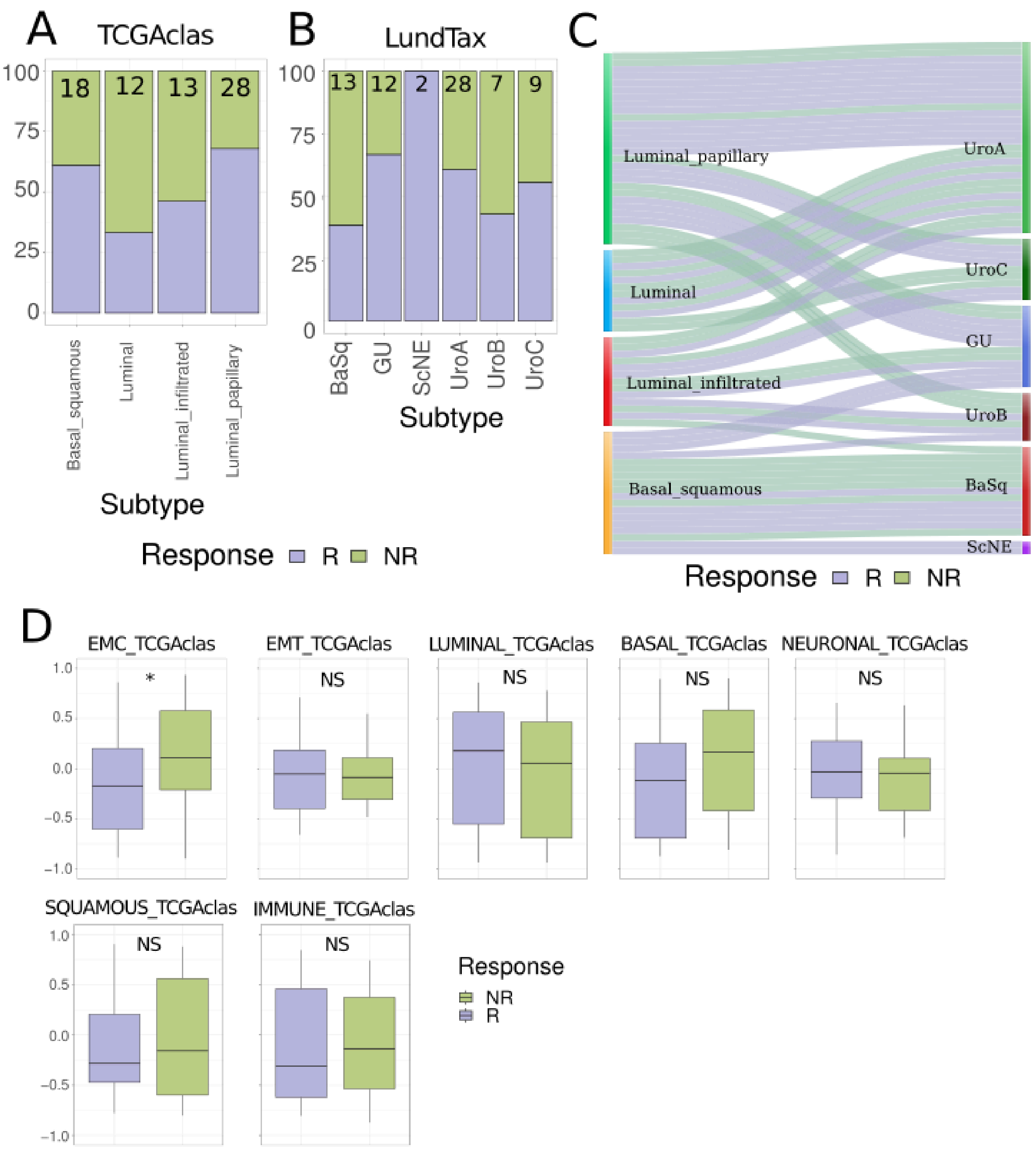


***Supplementary Figure 3. Molecular subtype classification results using TCGAclas and LundTax classifier A.*** *Proportions of the molecular subtype classification using TCGAclas classifier.* ***B.*** *Proportions of the molecular subtype classification using LundTax classifier.*
***D.*** *Sankey diagram illustrating the relationship between molecular subtype classifications assigned by the the TCGAclas classifier (left) and LundTax classifier (right). Each link represents a sample, showing how it is categorized by both classification methods. The color of the links corresponds to the response variable.* ***C.*** *TCGAclass signatures scores obtained by GSVA and its association with NAC response (EMT_TCGAclas, BASAL_TCGAclas, NEURONAL_TCGAclas, LUMINAL_TCGAclas, IMMUNE_TCGAclas, SQUAMOUS_TCGAclas p>0.05, EMC_TCGAclas p=0.029).*

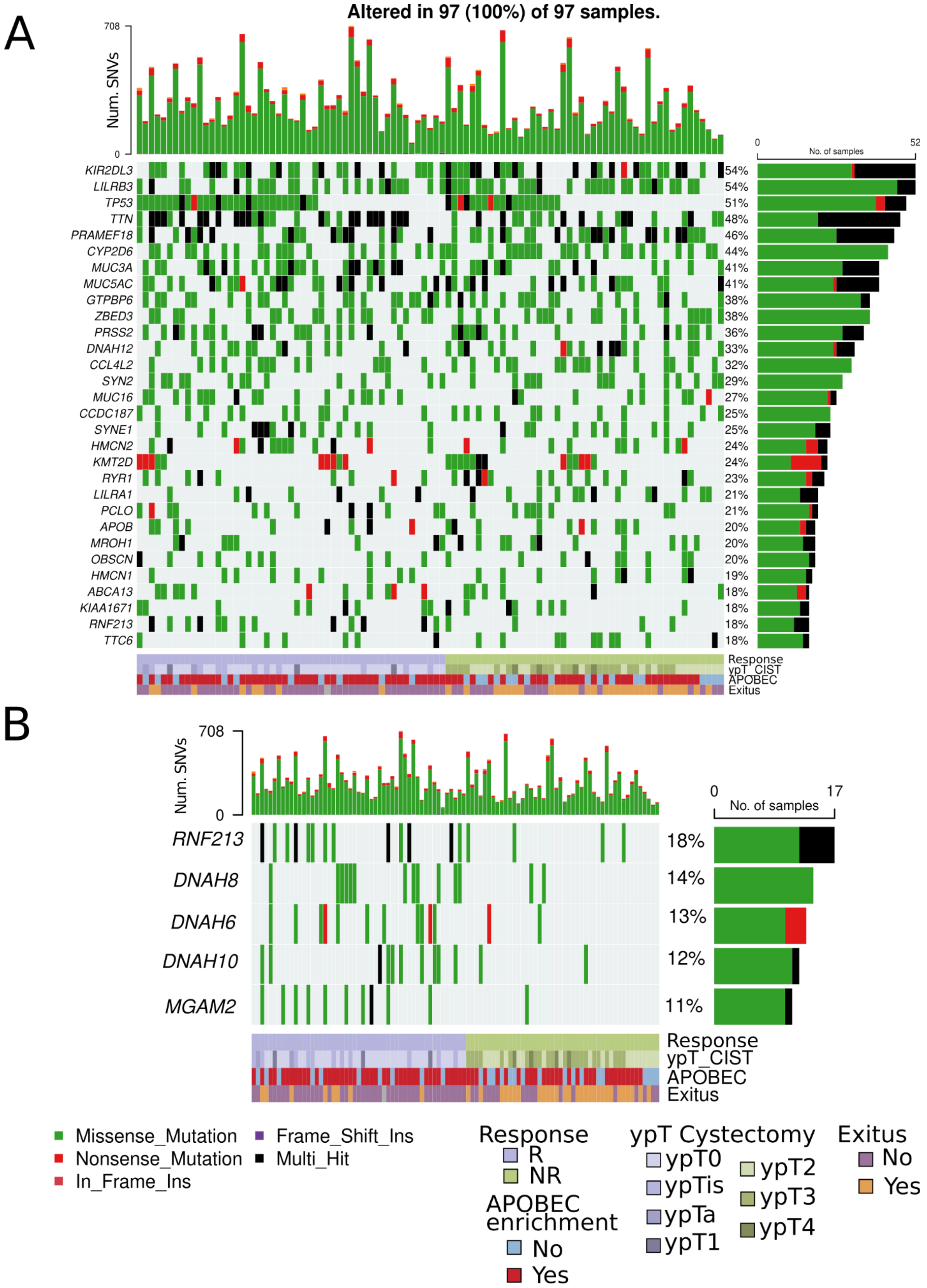


***Supplementary Figure 4. Oncoplot of top 30 mutated genes in our MIBC cohort and significantly different mutated genes. A.*** *Mutational landscape of top 30 mutated genes in our MIBC cohort with clinical and APOBEC information.* ***B.*** *Mutational landscape of genes*

*which were significantly different mutated in our MIBC cohort (MGAM2, DNAH8, DNAH6, DNAH10, RNF213) with clinical and APOBEC information.*

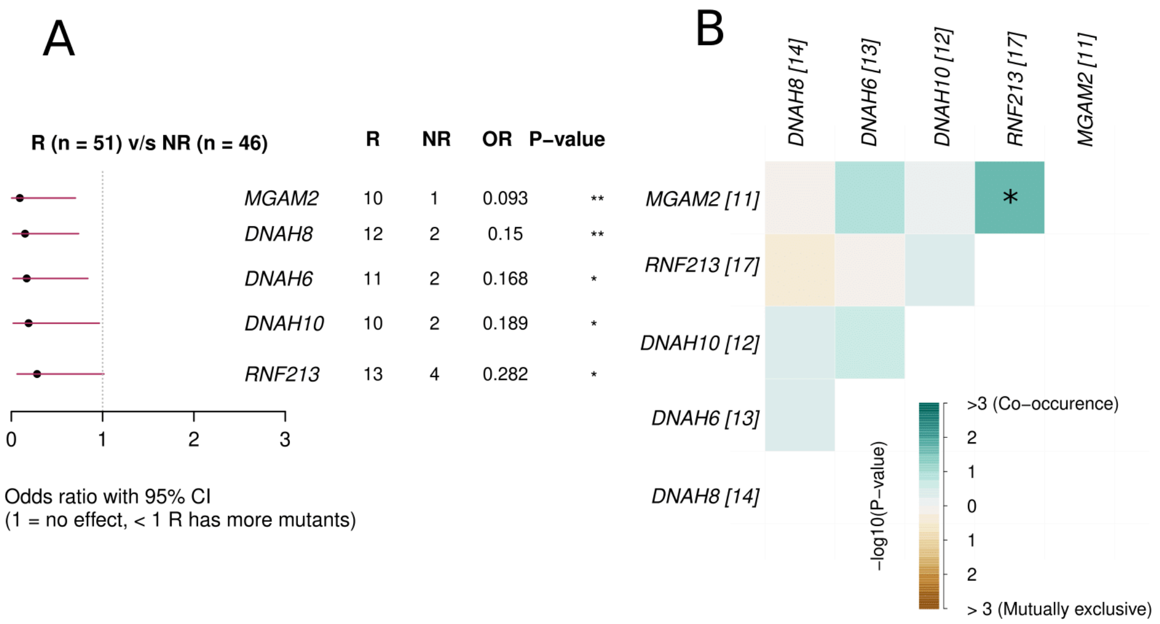


***Supplementary Figure 5. Somatic mutations associated with R and its co-occurrence. A.*** *Statistically significant differentially mutated genes between R and NR (MGAM2 p=0.0086, DNAH8 p=0.0086, DNAH6 p=0.0163, DNAH10 p=0.0301, RNF213 p=0.0349)* ***C.*** *Co-occurence of significant differentially mutated genes between R and NR. Green represents positive odds ratio and brown represents negative odds ratio.*

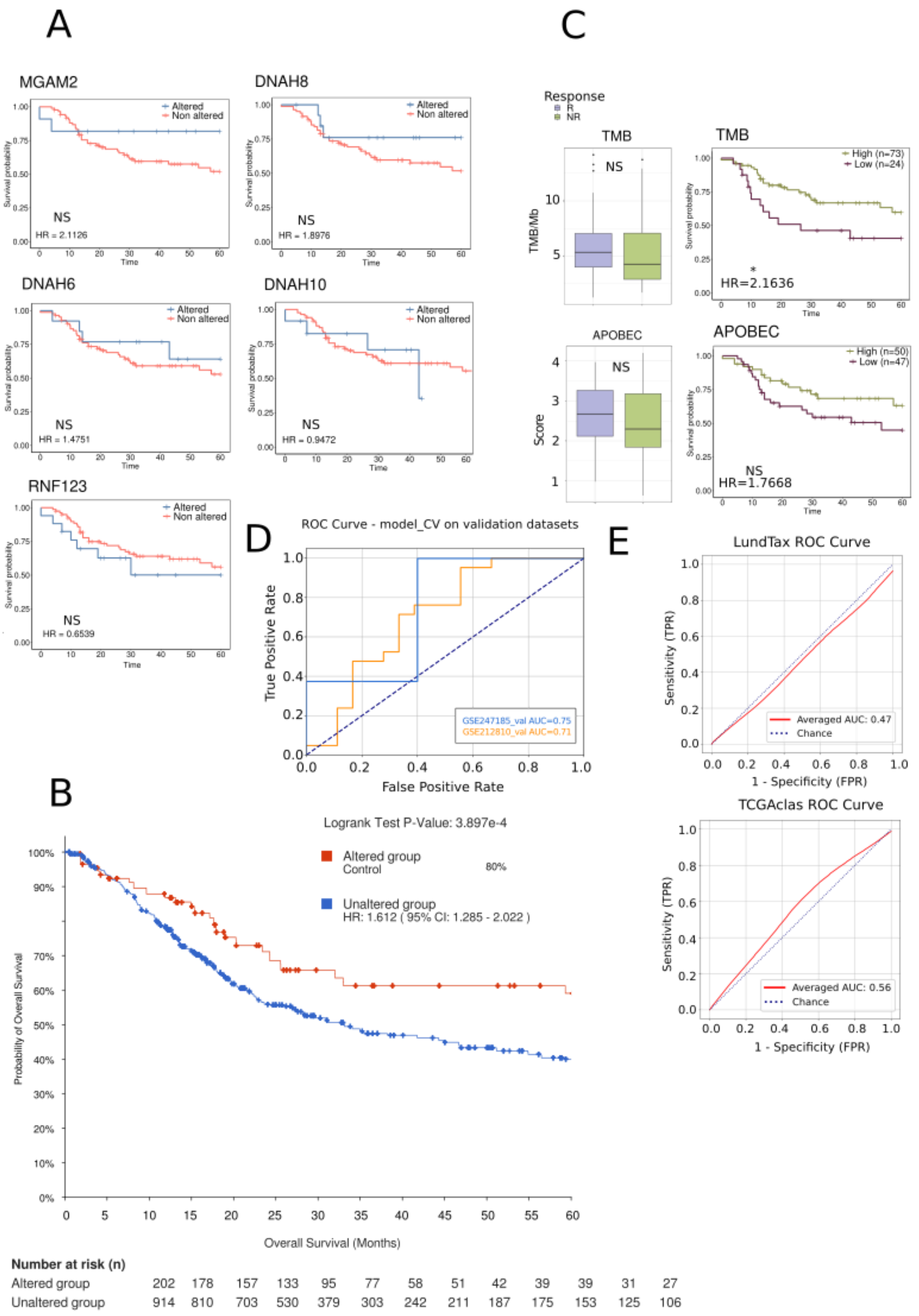


***Supplementary Figure 6. Kaplan Meier curves for significantly differentially mutated genes between R and NR, APOBEC and TMB association with response and ROC curves of WNT, LundTax and TCGAclas classifiers. A.*** *Kaplan-Meier curves showing the association between mutations in statistically significant differentially mutated genes and 5-year overall survival (OS) (p>0.05).* ***B.*** *Kaplan Meier curve of mutations in any of the DNAH genes family (DNAH8, DNAH6 and DNAH10) in cBioportal using several available*

*bladder cancer datasets (p=3.897e-4, HR=1.612, 95%CI=1.28-2.00).* ***C.*** *Association of tumor mutational burden (TMB) per Megabase (TMB/Mb) and APOBEC enrichment score with NAC response (p>0.05) and 5-year OS (TMB p=0.0231, HR=2.1636, 95%CI=1.09-4.28 ; APOBEC p>0.05)* ***D.*** *Roc curve resulting from the validation of the transcriptomic-based Xgboost (XGB) machine learning (ML) model (XGB-R) with two external and independent datasets (GSEXX: AUC=0.71 and GSEXX: AUC=0.75)* ***E.*** *ROC curves resulting from XGB models using only LundTax and TCGAclas classification results (LundTax AUC=0.47, TCGAclas=0.56).*
