## Supplementary material for "Transcriptomics-Driven Machine Learning Models Accurately Predict Chemotherapy Response in Muscle-invasive Bladder Cancer": Supplementary_Materials_and_Methods.pdf

### Patients and samples

A total of 130 patients with MIBC were retrospectively collected from four Catalan hospitals in Spain. Samples and data from patients included in this study were provided by the Hospital del Mar Biobank (MARBiobanc) and IGTP-HUGTP Biobank, both integrated in the Spanish National Biobanks and Biomodels Network of Instituto de Salud Carlos III (PT20/00023 and PT20/00050) and Tumor Bank Network of Catalonia. They were processed following standard operating procedures with the appropriate approval of the Ethical and Scientific Committees.

All patients received neoadjuvant chemotherapy (NAC) with either cisplatin-gemcitabine or dose dense MVAC (Methotrexate, Vinblastine, Adriamycin and Cisplatin). Patients achieving a dowstaging to non-MIBC status with no pathological lymph-node involvement (<=pT1N0) observed at cystectomy were defined as responders. For each patient, formalin-fixed paraffin-embedded (FFPE) pre-treatment samples were obtained after transurethral removal of bladder tumor (TURB). Out of the total 130 patients, 100 were eligible for molecular profiling. Among these, 100 pre-treatment samples were available to perform RNA-seq analysis (n=71) and WES analysis (n=97). A flow chart with details about samples and patients included is shown in Figure 1.

### Statistical analyses

Descriptive statistics of several clinical variables within a cohort of 100 patients, comparing between responders and non-responders, was performed using CompareGroups R package v.4.8.0 (method = 4) Kaplan–Meier survival curves were generated with survival v.3.4.0 and survminer v.0.4.9 R packages and p.values were obtained using a log-rank test.

### RNA and DNA sequencing

RNA extraction and library preparation was performed by the MARGenomics core facility at Hospital del Mar Institute for Medical Research. Samples were extracted using the AllPrep DNA/RNA FFPE kit from Qiagen following the manufacturer's instructions. RNA samples were degraded as expected due to the formalin and paraffin protocols to preserve tissue's histology. For RNA libraries, a combination of two Illumina protocols was utilized: TruSeq RNA Exome for the amplification steps (steps 1-6 of the published protocol), followed by the

RNA Prep with Enrichment protocol for pool exome enrichment (step 5 to 10, the last of the published protocol). For highly degraded FFPE samples, the best results were obtained by combining these two protocols rather than exclusively using the RNA Prep with Enrichment protocol. DNA libraries were prepared following the xGen cfDNA & FFPE DNA library Prep and xGen hybridization capture of DNA libraries, both from IDT. DNA and RNA libraries were enriched for coding regions, thereby improving sequencing data and optimizing read depth. Quality control of the libraries was conducted using the Bioanalyzer 2100 expert, ensuring that only samples with sufficient quantity were included in the sequencing step

Sequencing of DNA and RNA libraries was conducted at the Centre de Regulació Genòmica (CRG) core facility using NextSeq 2000, with 2x50 read length and an average of 25 million reads per sample. qPCR was performed in order to properly quantify the libraries

### RNA-Seq bioinformatics analyses

Raw fastq files were quality controlled using fastQC and fastqscreen. Subsequently, alignment was performed using STAR v.2.7.8 using GRCh38 genome as a reference and version 41 of hg38 GTF from gencode as annotation. Additionally, Picard v.2.25.1 was used to check the quality of the alignment. Quantification was performed using featureCounts from Subread package v.2.0.3 and version 41 of hg38 GTF from gencode as annotation. Minimum library size was 1.5 million counts. Lowly expressed genes (rowSums(counts.m>10) < 42) were removed for further analyses. Principal Component Analysis (PCA) and Hierarchical Clustering (HC) were performed in order to check and remove any outliers or groups due to either clinical or technical variables.

#### Differential expression analysis

Limma package v.3.54.2 was used to perform a differential expression analysis between responders and non-responders using TMM normalized counts obtained by edgeR v.3.40.2. Age, sex and batch were used to adjust the model as a fixed effect, while hospital variable was added to the model as a random effect. Voom function was used to model the mean-variance relationship. False discovery rate was used to correct for multiple testing. Due to the lack of significant adjusted p-values, genes were considered differentially expressed with p-value < 0.05 and |logFC| >1.

#### Molecular subtype classification

LundTax molecular subtype classification was performed using LundTax2023Classifier R package v.1.1.1 and log2TPM normalized counts. TCGAclas molecular subtype classification was also performed using BLCAsubtyping R package v.2.1.1 using log2TPMs. Differences in molecular subtype proportions between responders and non-responders were calculated using Fisher's exact test. GSVA R package v.1.46.0 was employed to perform a single-sample gene signature scoring of the different LundTax and TCGAclas classifiers' signatures and results were compared between R and NR using Wilcoxon test. In order to facilitate understanding the functionality of one of the most significantly different signatures between R and NR, we changed the name of Circuit by CELL CYCLE REG and ECM and smooth muscle by ECM. The nomenclature for KRT remained unchanged. Survival analysis for LundTax and TCGAclas signatures which are different between R and NR was also performed using survival v.3.4.0 and survminer v.0.4.9 R packages. Optimal cut-off point to separate high and low expression for the signatures scores was calculated using surv_cutpoint from survminer v.0.4.9 R package.

**Weighted gene correlation network analysis (WGCNA)**

Weighted gene correlation network analysis (WGCNA) using the R WGCNA package v1.72-5 was used to find clusters (modules) of highly correlated genes using the table of counts in TMM and setting the split number to 3 with a minimum cluster size of 25 genes. Pearson correlation was performed to calculate correlation between the first principal component of cluster genes and clinical variables. Over-representation analysis (ORA) was conducted on genes within the cluster that showed significant correlation with the response variable (green cluster, n=77), using the *enricher* function from clusterProfiler R package v.4.6.2 using an adjusted p-value <0.05 and Hallmark, C2 and C5 GO BP MSigDB collections v7.5.1. Genes related to WNT signaling pathway from C5 collection (GOBP_CANONICAL_WNT_SIGNALING_PATHWAY, GOBP_CELL_CELL_SIGNALING_BY_WNT, GOBP_REGULATION_OF_CANONICAL_WNT_SIGNALING_PATHWAY, GOBP_REGULATION_OF_WNT_SIGNALING_PATHWAY,GOBP_NEGATIVE_REGULATION_OF_CANONICAL_WNT_SIGNALING_PATHWAY, GOBP_NEGATIVE_REGULATION_OF_WNT_SIGNALING_PATHWAY) were put together and intersected with DE genes resulting into 17 genes.

**Top10up, Top10dn, WNT, EMC, KRT and CELL CYCLE REG gene expression signatures**

GSVA R package v.1.46.0 was employed to perform a single-sample gene signature scoring of the Top10up, Top10dn, WNT, EMC, KRT and CELL CYCLE REG gene expression signatures (Supplementary Table 9). Signature scores were calculated using the log2(TPM) table of counts and GSVA v.1.46.0. Signature scores were compared between R and NR using the Wilcoxon rank sum test. Optimal cut-off point to separate high and low expression for the signature scores was calculated using surv_cutpoint from survminer v.0.4.9 R package. Survival analysis was also performed as previously described.

A heatmap was performed with the expression of the five signature genes (Top10up, Top10dn, WNT, EMC, KRT and CELL CYCLE REG) using the score obtained by GSVA R package v.1.46.0 to detect any cluster based on the variable response. Correlation among genes from all identified signatures in the RNA-Seq data was performed to avoid the multicollinearity, enhancing the robustness and stability of the future ML algorithm.

### Whole exome sequencing (WES) bioinformatics analyses

Fastq files were processed with the nf-core/sarek pipeline v.3.4.0 with default parameters and reference genome for GRCh38. Raw fastq files were quality controlled using fastQC v.0.12.1 and fastp v.0.23.4. Subsequently, an alignment step was performed using BWA 0.7.17 and its quality was checked using Picard v.2.25.1. Variant calling (VC) was performed using mutect2 from GATK4 v.4.4.0.0 in tumor-only mode. Additionally, the quality of the identified variants was assessed using the FilterMutectCalls function. VCF files were annotated using VEP v.10.2.0 and transformed to MAF objects using vcf2maf v.1.6.21. Only variants with quality filter “PASS” were included in downstream analyses. Additionally, we removed variants annotated in gnomAD v.2.1.1 or exhibiting a gnomAD allele frequency (AF) > 0.01. We also filtered out variants with low sequencing depth (DP < 20) and those with AF > 0.95 or AF < 0.05.

#### Somatic mutation landscape

MAFtools R package v.2.18.1 was used to analyze somatic mutations from MAF files. The top 15 bladder cancer mutated genes (Supplementary Table 5) were plotted using oncoplot function. Additionally, TMB was calculated using the tmb function. TMB from our cohort was compared to the TCGAclas-BLCA cohort using the TCGAclasCompare function. The APOBEC score was calculated using the trinucleotideMatrix function. Moreover, the function mafCompare was used to detect significantly different mutated genes between R and NR. Differences in TMB and APOBEC score across the groups were assessed using the Wilcoxon rank sum test. Survival analysis was also performed using survival v.3.4.0 and survminer v.0.4.9 R packages separating patients by mutated and non-mutated genes. Moreover, in order to validate the importance of DNAH alterations, a survival analysis was performed using cBioPortal for Cancer Genomics (https://www.cbioportal.org/). We selected those patients in any of the following datasets: MSK Eur Urol 2014, MSK J Clin Oncol 2013, MSK Nat Genet 2016, MSK/TCGA 2020, TCGA Cell 2017, BGI Nat Genet 2013, DFCI/MSK Cancer Discov 2014, TCGA PanCancer Atlas, BCAN/HCRN Nat Commun 2022, and Cornell/Trento Nat Genet 2016. Patients with mutations in DNAH8, DNAH6 and DNAH10 were compared against patients without mutations in those genes.

#### SCNV

Somatic copy number variation (SCNVs) analysis was conducted using CNVkit v.0.09.10 in tumor-only mode. For each sample, .cns files obtained by CNVKit were merged in a GenomicRanges object splitted by sample using GenomicRanges R package v.1.50.2. Segmented log2 copy number ratios were transformed into the following states: 0, 2-copy loss; 1, 1-copy loss; 2: normal; 3, 1-copy gain; 4, >= 2-copy gain. Genes related to bladder cancer were selected and plotted using cnvOncoPrint function from CNVRanger R package v.1.14.0 to get the landscape of amplifications and deletions. Important genes previously reported in the literature were also chosen to perform a statistical analysis to detect differences between R and NR in indels and amplifications using CompareGroups R package v.4.8.0 (method = 4). Survival analysis was also performed using survival v.3.4.0 and survminer v.0.4.9 R packages separating patients by the presence of deletions or amplifications.

### Predictive models of response to NAC

The machine learning framework developed for this project consists of two different steps. The first one encompasses all preprocessing steps, including scaling and splitting data. During the training phase, optimal hyperparameters for the ML algorithm are determined through 15-fold cross-validation, repeating the process multiple times to ensure the robustness and avoid overfitting for a specific seed. Subsequently, the model is evaluated using the traditional train/test split method, where 70% of our data is used for training and the remaining 30% is reserved for testing. Moreover, bootstrapping was also used to reduce overfitting and improve the accuracy.

We selected a set of variables significantly associated with R, ensuring low correlation among themselves and avoiding the overlap in case of gene signatures. The final model (XGB-RW) includes the following variables: Top10up, Top10dn, WNT, EMC, KRT and CELL CYCLE REG signatures (GSVA), alterations in DNAH8, DNAH6 or DNAH10 (0=non-altered, 1 altered), deletions in KDM6A (2=no deletion, 1=deletion).

Finally, a Xgboost (XGB) model was used for the final model. XGB was chosen because it is very robust with overfitting. Additionally, to mitigate the consequences of our limited sample size, we tested the model using 1000 different seeds to achieve more reliable and consistent scores. Moreover, the Bootstrap .632+ method was applied as an internal validation strategy, providing information about how the model might perform on datasets not included in the training phase. +++ We have also run the model using different features in order to know whether the AUC is stable or not. We tested models using: only RNA-Seq features (XGB-R), only WES features (XGB-W), both RNA-Seq and WES features (XGB-RW) and using only the WNT (XGB-WNT) score.

The ROC curve shown in the figures represents an interpolation based on the mean AUC obtained from 1,000 independent runs of model training and validation, ensuring a robust and representative assessment of performance.

To assess the generalizability of the XGB-R model, two publicly available transcriptomic datasets from the Gene Expression Omnibus (GEO) database were used for external validation: GSE237185 (n = 13) and GSE212810 (n = 39).
